# Impaired cortical gap-mediated inhibition in chronic tinnitus but with limited individual diagnostic accuracy

**DOI:** 10.64898/2026.11.11.26367707

**Authors:** Hongyu Lu, Payam S. Shabestari, Milena Engelke, Shagun Ajmera, Elva Arulchelvan, Christoph Braun, Inés S. Fernández del Campo, Namitha Jain, Emilia Kaniewska, Tobias Kleinjung, Johann Korn, Jan Ost, Lauren Ralston Ruesch, Sarah Rambour, Nathan Shields, Susanne Staudinger, Beat Tödtli, Stephan M. Wolpert, Anusha Yasoda-Mohan, Fatima T. Husain, Julia Campbell, Dirk De Ridder, Berthold Langguth, Winfried Schlee, Sven Vanneste, Patrick K.A. Neff, Christopher R. Cederroth

## Abstract

Tinnitus lacks an objective marker, limiting diagnosis and treatment evaluation. We tested whether gap-mediated inhibition of cortical sound-evoked responses provides one. In a prospective, multicentre case–control study at seven sites in the United States and Europe, adults with and without chronic tinnitus heard brief pulses preceded or not by a silent gap, embedded in broadband or narrowband carriers, during electroencephalography. The primary outcome was inhibition of the N100 response. Of 584 eligible participants, 455 yielded high-quality recordings with complete covariate data (232 cases, 223 controls; mean age 45.4 years, 52% women) that entered the adjusted analyses. Inhibition was strongest for broadband noise whereas narrowband carriers gave limited dynamic range. Participants with tinnitus showed reduced broadband gap-mediated inhibition (β = −19.27%, d = −0.413, cluster-corrected p = 0.002), scaling with tinnitus severity (covariate-adjusted √THI model: partial R^2^ = 0.049, p = 0.0015). In contrast, discrimination of individuals was limited (unadjusted AUC 0.610, 95% CI 0.559–0.660). Cortical gap inhibition is a reproducible group-level effect rather than an individual diagnostic test.

**Study Registration:** NCT06520865

## Introduction

Subjective tinnitus is a phantom percept whereby an individual perceives sound(s) in absence of an external source^1,2^. Similar to the phenomenon of phantom limb percept, tinnitus is thought to emerge as a central maladaptation to missing peripheral input from the ear^3^, such as sensorineural or conductive hearing loss – the two major risk factors for tinnitus^4^. Despite a global prevalence of 14.4%^5,6^, effective treatments are still lacking, solely relying on the alleviation of the psychological burden by cognitive behavioral therapy. Recent large interventional clinical trials suggest that bimodal modulation, based on the combined stimulation of auditory and somatosensory routes, can diminish tinnitus distress^7–11^. However, all these studies rely on self-report questionnaires with responses that may vary from day to day depending on the psychological status of the patient. The establishment of objective, quantitative, and translationally valid biomarkers for tinnitus severity would provide a considerable advance for the reliable assessment of interventional trials on tinnitus.

Biomarker discovery in the tinnitus field has been challenging, with often inconclusive results, and relying on small sample sizes^12^. Recent progress includes a large analysis from the Nurses’ Health Study (NHS), which found several plasma metabolites associated with persistent tinnitus^13^. In contrast, a high sensitivity proximity elongation assay failed in detecting plasma inflammatory biomarkers in subjects with constant tinnitus (N= 1622)^14^. However, in a subset of these participants (N=405), auditory brainstem responses (ABR) revealed delayed Wave V latencies in individuals with constant tinnitus, a result replicated in an independent study^15,16^. Recently, Smith et al. used an innovative approach based on pupil dilations and facial movements elicited by emotionally evocative sounds to accurately measure tinnitus distress and hyperacusis^17^.

Promising pre-clinical research in the tinnitus field has investigated an objective behavioral test based on the suppression of the startle response by the presence of a preceding silent gap embedded in a carrier background noise, the so-called Gap Pre-pulse Inhibition of the Acoustic Startle (GPIAS)^18,19^. Validated against conditioning behavioral tests and supported by numerous neural correlates of tinnitus^20–22^, GPIAS has been used to demonstrate the efficacy of bimodal stimulation in rodents prior to testing in humans^10,23^. Its translation to humans has, however, been controversial, with poor reliability of the blinking response^24^, and misconceptions around the conscious perception of the gap as well as the reflex mechanism driving the suppression of the startle response^25,26^.

With increasing evidence that GPIAS is regulated at the level of the auditory cortex^27–29^, electroencephalography (EEG) approaches have been explored as a means to examine cortical analogues of the GPIAS^30–32^. Magnetoencephalography (MEG) shows even greater sensitivity than EEG or blinking responses with evoked-responses to sound pulses as high as 90 dB SPL being suppressed by 72.5% in presence of a gap in healthy, normal hearing participants.^33^ Because electroencephalography (EEG) is much more amenable to clinical routine, we sought to test the ability of this paradigm to inhibit pulse-elicited event-related potentials (ERPs) in the presence of a gap. Here, we aimed to test this approach as a potential biomarker for the individual diagnosis of tinnitus. For this purpose, we investigated a large sample of participants at 7 sites, allowing for a high-powered aggregated sample.

## RESULTS

This study assesses the adaptation of the behavioral GPIAS paradigm to cortical measures using EEG. The cortical-GPIAS (cGPIAS) paradigm consisted of presenting a 20 ms white noise pulse at 90 dB SPL embedded in a continuous carrier of background noise. This embedded pulse elicits a distinguishable ERP with a pronounced N100 deflection. The addition of a 50 ms silent gap 240 ms before the pulse is hypothesized to lead to pre pulse inhibition resulting in a lower amplitude of the ERP (**Figure 1a**). Sessions with three different carrier sounds [broadband noise (BBN), 3 kHz-centered narrowband noise (NBN), and 8 kHz-centered NBN)] were randomly presented and contained 100 trials of pulse only (PO), gap and pulse (GP) or gap only (GO) conditions, the latter being used as an internal control to verify the absence of ERPs when a pulse would have otherwise occurred. Additional twenty-five PO trials of varying intensity (70, 75, 80, 85, 90 dB SPL) were presented before and after the three core sessions to assess habituation (**Figure 1b**). Of 820 individuals assessed for eligibility, 584 were enrolled between September 2024 and October 2025, and a complete record across questionnaires, audiometry and EEG was available for 562 of them (Figure 1c, Table S3). The most frequent participant exclusions were due to intermittent tinnitus or severe hearing loss. Participants were 18 to 70 years old [mean age of 45.4 (SD ± 16.1)] and 52% were female. The majority was highly educated (72.8% University level), and none were cochlear implant/hearing aid users (Table 1). No adverse events related to the paradigm were reported; participant comfort and sleepiness were checked via intercom after each block.

**Figure 1.**
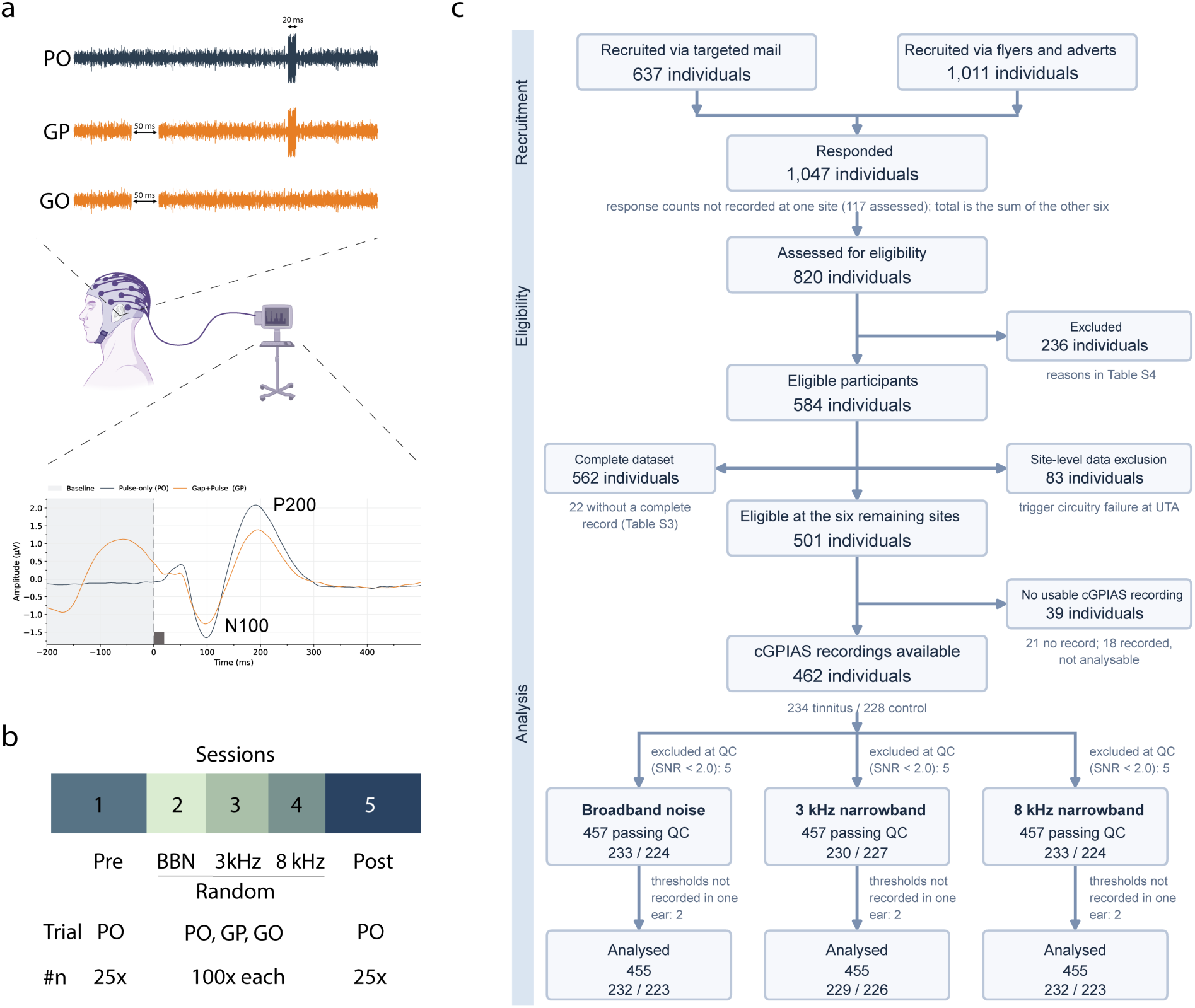
Study design and recruitment flowchart. (a) Schematic illustration of the gap prepulse inhibition of the acoustic startle (GPIAS) paradigm and corresponding EEG response. Trials were presented on a continuous noise carrier and consisted of pulse only (PO), gap and pulse (GP), and gap only (GO) conditions (used as an internal control). The pulse duration was 20 ms and the gap duration was 50 ms, interspaced by 240 ms interstimulus interval. The lower panel shows representative PO and GP event-related potentials (ERPs), illustrating the difference in the evoked response following pulse onset (black box) in presence (orange) or absence (black) of a gap. N100 and P200 peaks are shown. (b) Experimental session structure consisting of Sessions 1 (beginning) and 5 (at the end) with PO only trials, with 25 repetitions of randomly presented pulse stimuli ranging from 70 dB to 90 dB. Sessions 2 to 4 consisted of BBN, 3 kHz, and 8 kHz carriers presented in randomized order, with PO, GP, and GO trials of 100 repetitions each. (c) Participant flow through recruitment, eligibility assessment, and GPIAS quality control. Final participant numbers contributing to the N100 analyses are shown separately for the BBN, 3 kHz, and 8 kHz carrier conditions. Quality control was applied separately to each carrier, so the three analysed samples differ in composition: 449 participants passed all three carriers and 461 passed at least one. Cross-validation analyses re-derive cluster selection and the outlier fence within training samples, so of the 455 participants analysed in the broadband carrier, 447 contribute to the leave-one-site-out and 454 to the five-fold estimates.

**Table 1.**
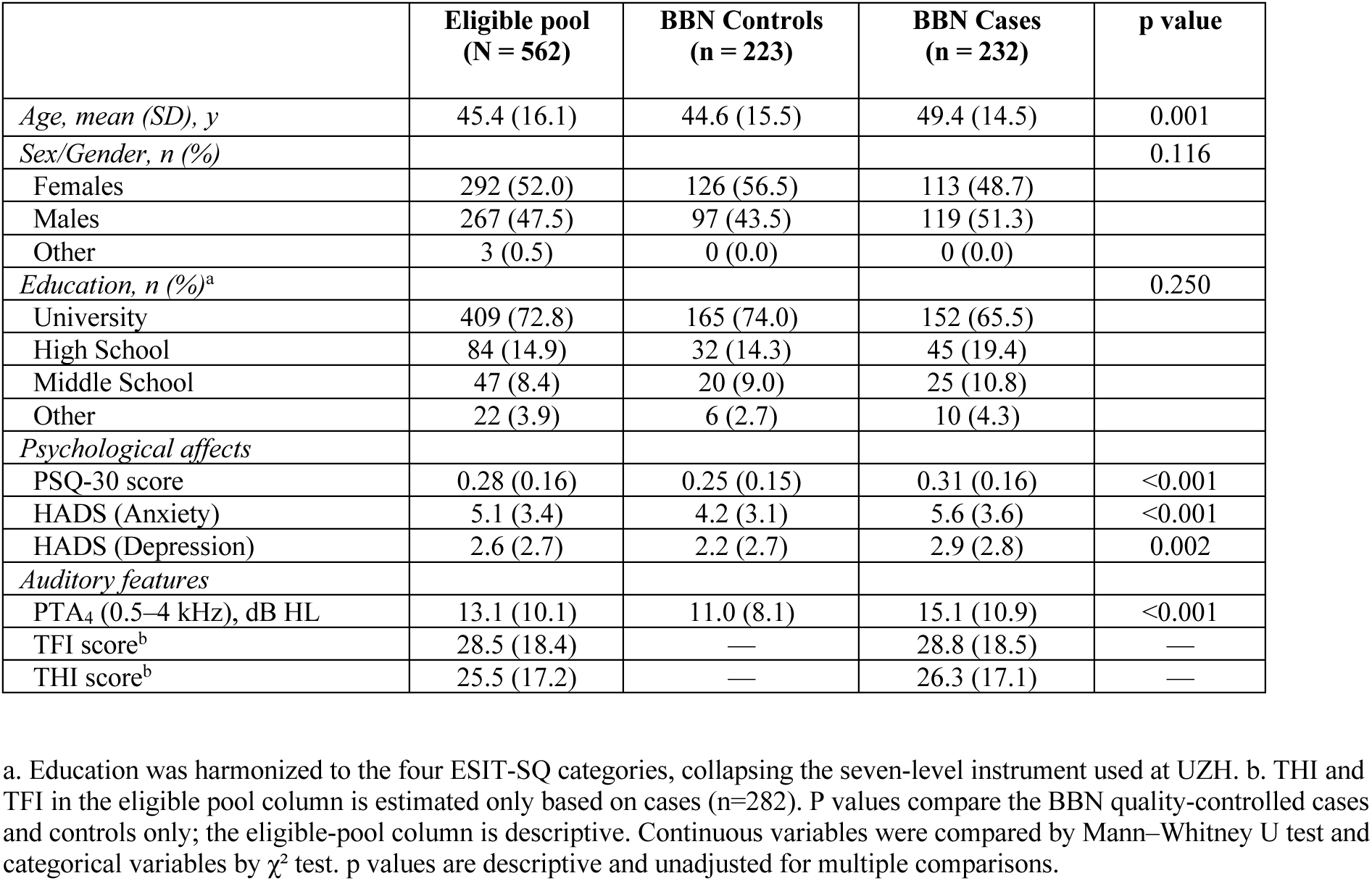
Characteristics of the TIDE participants with a complete record (N = 562)

|  | **Eligible pool (N = 562)** | **BBN Controls (n = 223)** | **BBN Cases (n = 232)** | **p value** |
|---|---|---|---|---|
| *Age, mean (SD), y* | 45.4 (16.1) | 44.6 (15.5) | 49.4 (14.5) | 0.001 |
| *Sex/Gender, n (%)* |  |  |  | 0.116 |
| Females | 292 (52.0) | 126 (56.5) | 113 (48.7) |  |
| Males | 267 (47.5) | 97 (43.5) | 119 (51.3) |  |
| Other | 3 (0.5) | 0 (0.0) | 0 (0.0) |  |
| *Education, n (%)*<sup>a</sup> |  |  |  | 0.250 |
| University | 409 (72.8) | 165 (74.0) | 152 (65.5) |  |
| High School | 84 (14.9) | 32 (14.3) | 45 (19.4) |  |
| Middle School | 47 (8.4) | 20 (9.0) | 25 (10.8) |  |
| Other | 22 (3.9) | 6 (2.7) | 10 (4.3) |  |
| *Psychological affects* |  |  |  |  |
| PSQ-30 score | 0.28 (0.16) | 0.25 (0.15) | 0.31 (0.16) | <0.001 |
| HADS (Anxiety) | 5.1 (3.4) | 4.2 (3.1) | 5.6 (3.6) | <0.001 |
| HADS (Depression) | 2.6 (2.7) | 2.2 (2.7) | 2.9 (2.8) | 0.002 |
| *Auditory features* |  |  |  |  |
| PTA<sub>4</sub> (0.5–4 kHz), dB HL | 13.1 (10.1) | 11.0 (8.1) | 15.1 (10.9) | <0.001 |
| TFI score<sup>b</sup> | 28.5 (18.4) | — | 28.8 (18.5) | — |
| THI score<sup>b</sup> | 25.5 (17.2) | — | 26.3 (17.1) | — |

a. Education was harmonized to the four ESIT-SQ categories, collapsing the seven-level instrument used at UZH. b. THI and TFI in the eligible pool column is estimated only based on cases (n=282). P values compare the BBN quality-controlled cases and controls only; the eligible-pool column is descriptive. Continuous variables were compared by Mann–Whitney U test and categorical variables by χ<sup>2</sup> test. p values are descriptive and unadjusted for multiple comparisons.

### Silent gaps suppress pulse-evoked responses

To determine whether EEG is sufficiently sensitive to capture gap-mediated inhibition of cortical evoked responses to sound pulses, we inspected grand averages and topographies for morphology, latency, and signal-to-noise ratio of N100 responses for each carrier and condition at Cz. During this examination, we identified a software failure in running GP trials at the University of Texas at Austin (UTA) site, which led us to exclude all cGPIAS data collected in UTA (n = 83).

From the remaining 6 sites, N100 amplitude at Cz increased monotonically with pulse levels, confirming an expected input/output function (**Figure 2a,b**). When comparing pre- and post-sessions (session 1 and 5, respectively), the N100 amplitudes in session 5 were systematically attenuated relative to those in session 1 (by 30% at 90 dB SPL, *p* < 0.001), reflecting moderate neural habituation over the 60-minute experiment. Importantly, N100 amplitudes remained robust and well above baseline even after habituation, confirming that the cGPIAS paradigm maintained sufficient SNR throughout the full experimental duration (**Figure 2b**).

**Figure 2.**
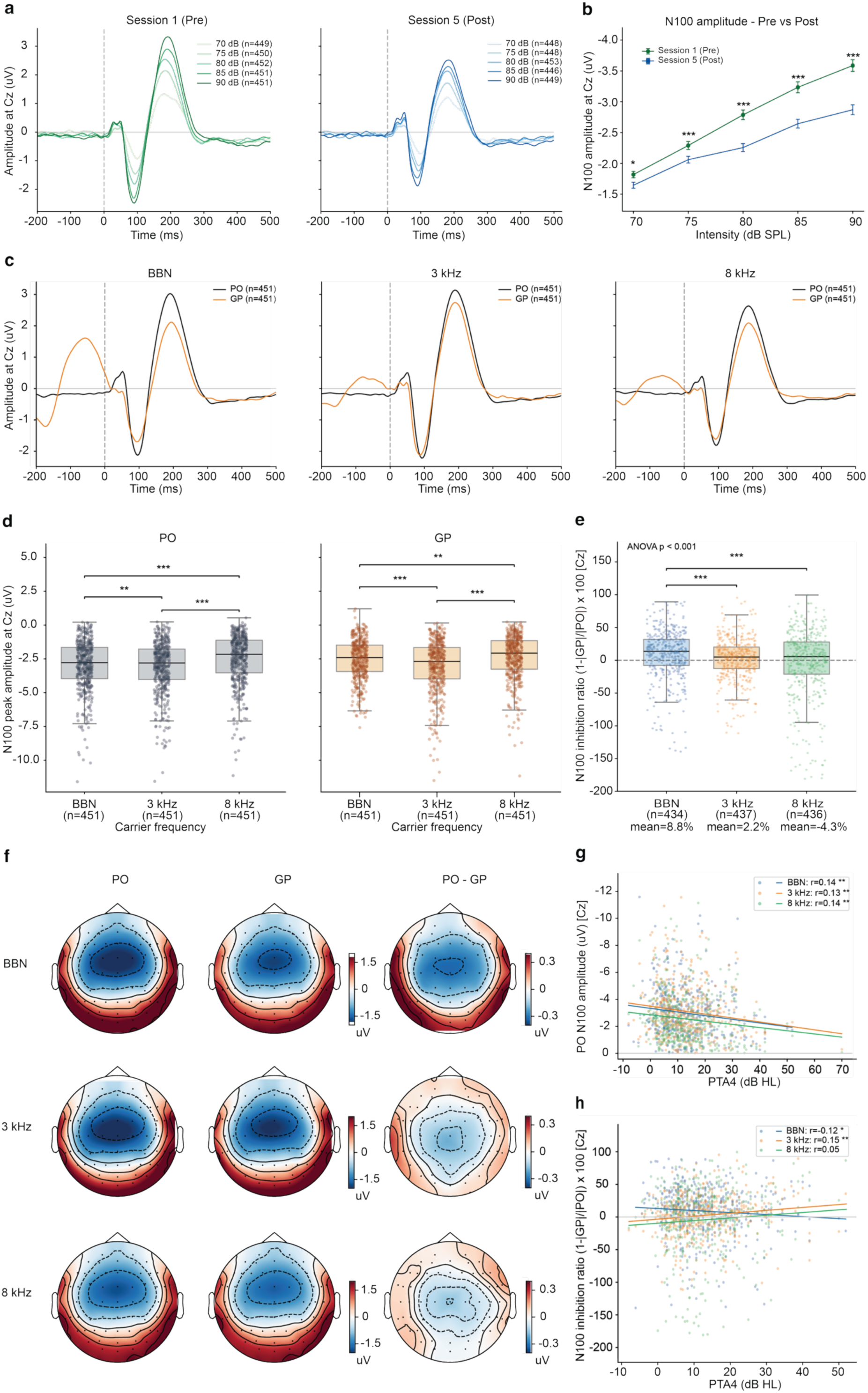
EEG can detect effectively gap-inhibited cortical sound-evoked responses at Cz in a broadband background. (a) Grand-average Cz ERPs for Session 1 (pre-exposure) and Session 5 (post-exposure), shown across pulse intensities from 70 to 90 dB SPL. (b) Mean N100 peak amplitude at Cz before and after exposure as a function of pulse intensity. Error bars indicate the standard error of the mean (SEM). (c) Grand-average PO (black) and GP (orange) waveforms at Cz for the BBN, 3 kHz, and 8 kHz NBN carrier conditions. (d) N100 peak amplitudes at Cz for PO (grey) and GP (light orange) by carrier. (e) N100 inhibition ratio at Cz for BBN (blue), 3 kHz (orange) and 8 kHz (green) carriers, calculated as (1-|GP|/|PO|) × 100. (f) Scalp topographies of the mean N100 PO response, GP response, and PO−GP difference over 80–120 ms for each carrier. Color bars indicate amplitude in µV. (g) Relationship between PO N100 peak amplitude at Cz and PTA_4_ hearing thresholds for BBN (blue), 3 kHz (orange) and 8 kHz (green) carriers. Lines indicate ordinary least-squares (OLS) fits, and annotations report Spearman correlation coefficients. (h) Relationship between the N100 inhibition ratio at Cz and PTA_4_ hearing threshold for each carrier. Lines and annotations are as described for panel (g). For (d) and (e), each dot represents an individual participant, with the distributions summarized in boxplots. Significance was assessed using the Friedman test, followed by pairwise Wilcoxon signed-rank tests. Benjamini–Hochberg false-discovery rate (FDR) was used to correct for multiple comparisons. Asterisks indicate statistical significance: p < 0.05 (*), p < 0.01 (**), and p < 0.001 (***).

When comparing the PO and GP responses at Cz (**Figure 2c**), we confirmed that cGPIAS can elicit neural inhibition across all carrier conditions, although to a lesser extent in 3 and 8 kHz NBN carriers as compared to BBN carrier (ANOVA, *p* < 0.0001, **Figure 2d**). The magnitude of the inhibition occurring during the BBN carrier measured using EEG (10.5%) was however strikingly lower than what we previously reported with MEG (72.5%), thus offering a smaller dynamic range to assess the presence of tinnitus, inferred by a reduced inhibition. Amplitude of the N100 response in PO and GP trials varied depending on the carrier environment (**Figure 2e**). This amplitude also varied only slightly at each site (**Figure S1**), possibly due to the use of different EEG hardware systems (**Table S1**). We believe that the smaller number of cortical neurons recruited during NBN carriers (**Figure 2c**) is insufficient to trigger an inhibition as effective as in the BBN carrier (**Figure 2d, e**). Topoplots confirmed the expected fronto-central distribution of N100 for PO and GP, typical of acoustically evoked ERPs (**Figure 1f, Figure S2**). However, paired permutations tests comparing PO vs GP within subjects show almost null patterns for NBN carriers, something also evident at each site (**Figure S2**).

The amplitude of the PO responses correlated with both ear’s pure-tone average audiometric thresholds at 4 frequencies (PTA_4_: 0.5, 1, 2, 4 kHz), regardless of the carrier frequency used (r = -0.13 to -0.14, *p* < 0.01, higher PTA_4_ thresholds were associated with smaller absolute amplitudes; **Figure 2g**). In contrast, while the degree of inhibition inversely correlated with PTA_4_ in BBN carriers (higher PTA_4_ thresholds were associated with less inhibition, the correlation was positive in 3 kHz and 8 kHz NBN carriers (although only significant for 3 kHz) (**Figure 2h)**. The correlational results with PTA_4_ were used to correct for hearing loss in the follow-up analyses.

Overall, we gathered high-quality BBN cGPIAS data across 6 sites in order to assess whether gap-mediated inhibition of pulse-ERP is impacted in individuals with tinnitus.

### Primary analysis: decreased inhibition in participants with tinnitus

Following quality control, 232 cases and 223 controls were analyzed for the BBN carrier condition. The two groups differed in age, in psychological features and in hearing thresholds, but not in sex or education (**Table 1**).

Conventional ordinary least squares (OLS) Freedman–Lane spatial cluster-permutation testing, with age, gender, PTA_4_, and site included in the regression model, identified reduced BBN N100 inhibition in tinnitus participants compared with controls in the full cohort. The effect involved an 11-electrode frontocentral cluster comprising AF3, AF4, AFz, C3, F1, F3, F4, F6, FC1, FC3, and Fz (cluster mass = 27.80; FWER-corrected permutation *p* = 0.0020; **Figure 3a,b**). The descriptive adjusted group difference across the cluster was −19.27 percentage points (d = −0.413). Site-specific estimates were negative at all six recording sites (**Figure 3c**), and this direction was maintained when each site was excluded in turn while retaining the 11-electrode cluster (β range, −22.54 to −13.54 percentage points; d range, −0.484 to −0.308; all FDR-corrected q ≤ 0.0018; **Figure 3d**)

**Figure 3.**
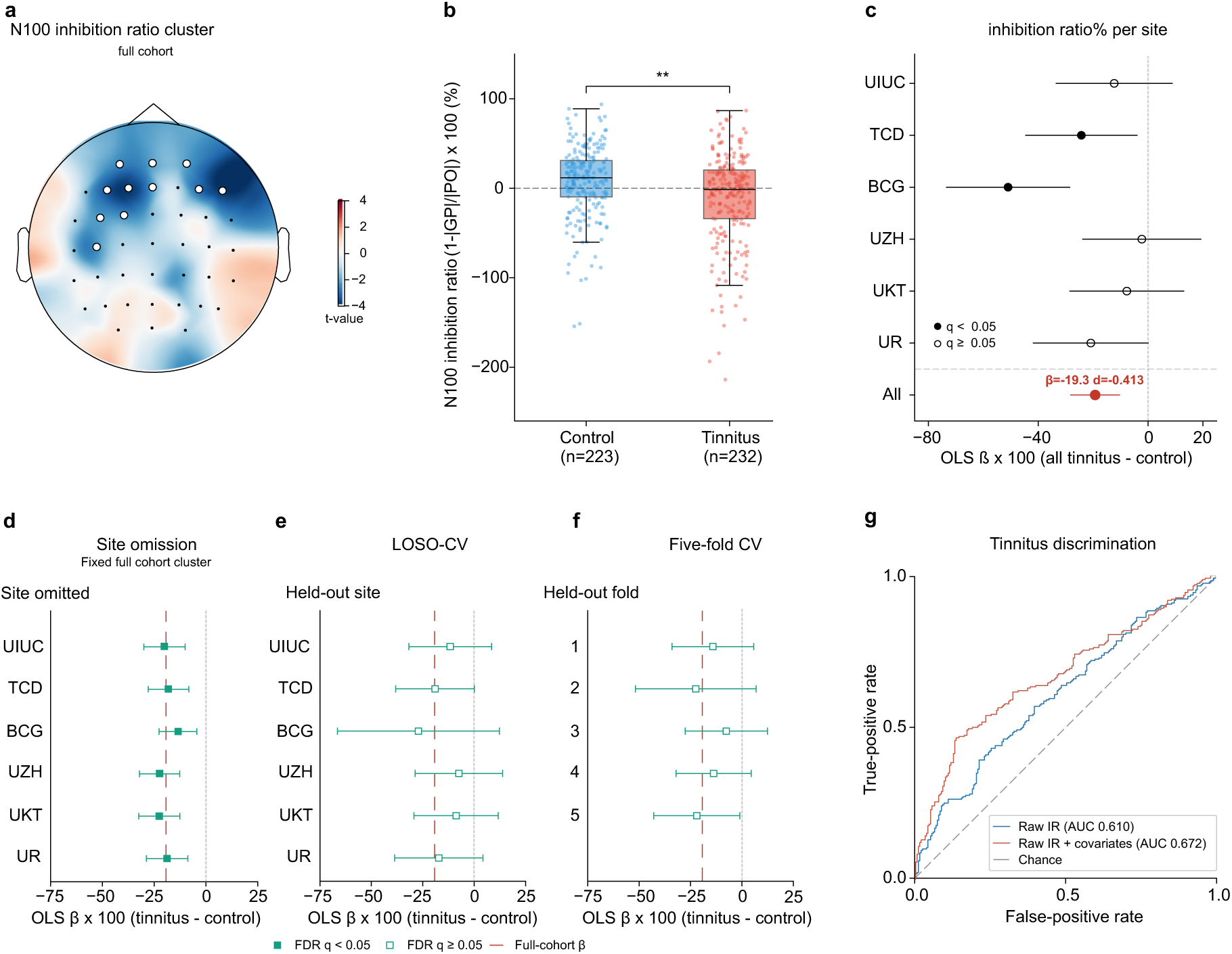
Data-driven EEG analysis reveals impaired inhibition in tinnitus. (a) Scalp map of t statistics for the tinnitus and control difference in BBN N100 inhibition ratio. White circles mark the 11-electrode cluster identified by covariate-adjusted (for age, gender, PTA_4_, and site) cluster-permutation testing in the full cohort (AF3, AF4, AFz, C3, F1, F3, F4, F6, FC1, FC3, and Fz). The color scale represents the t values. (b) Covariate-adjusted participant inhibition ratios averaged across eligible cluster electrodes. Blue and red indicate controls and tinnitus participants, respectively. Points represent individual observations and boxplots summarize the group distributions. The bracket and asterisks indicate spatially corrected cluster-permutation significance (** p < 0.01). (c) Site-specific and pooled (red) adjusted OLS estimates of the tinnitus minus control difference in BBN N100 inhibition ratio. Horizontal lines represent 95% confidence intervals. Filled site-specific markers indicate BH-FDR q < 0.05 across the six site comparisons; open markers indicate q ≥ 0.05. (d) Site omission analysis, excluding each site in turn while retaining the full-cohort cluster. (e,f) Held-out estimates from LOSO-CV and five-fold CV, respectively, using training-derived N100 windows, cluster selection, and nuisance adjustment. Forest plots show β in percentage points and two-sided 95% OLS confidence intervals. Filled markers indicate BH-FDR q < 0.05 within each panel’s comparison family; open markers indicate q ≥ 0.05. Red dashed lines in panels d–f indicate the full-cohort coefficient. (g) ROC curves for raw IR and IR combined with covariates (age, gender, PTA_4_, and site).

Leave-one-site-out cross-validation (LOSO-CV) analyses used training-selected clusters and training-derived nuisance adjustments to evaluate each held-out site. In the full cohort, adjusted differences were negative at all six held-out sites (β range, −27.11 to −7.47 percentage points; d range, −0.437 to −0.165), although none survived FDR correction (q range, 0.1535–0.2430; **Figure 3e**). Five-fold cross-validation (five-fold CV) estimates were likewise negative in every fold (β range, −22.43 to −7.62 percentage points; d range, −0.437 to −0.159), with no fold surviving FDR correction (q range, 0.1001–0.2252; **Figure 3f**; **Table 2**).

**Table 2.**
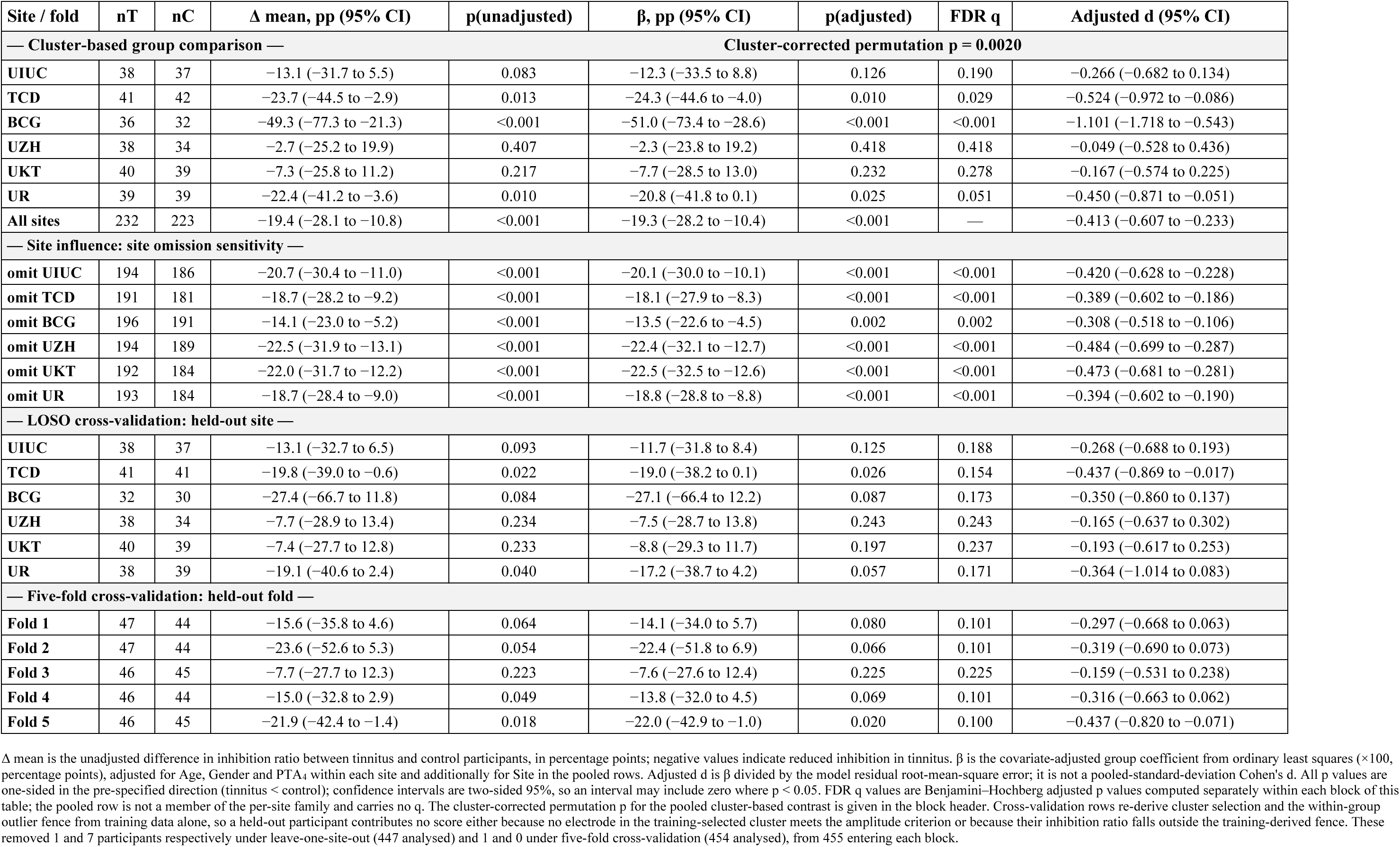
Group difference in N100 inhibition in the broadband noise carrier — full cohort.

| **Site / fold** | **nT** | **nC** | **Δ mean, pp (95% CI)** | **p(unadjusted)** | **β, pp (95% CI)** | **p(adjusted)** | **FDR q** | **Adjusted d (95% CI)** |
|---|---|---|---|---|---|---|---|---|
| **— Cluster-based group comparison —** |  |  |  | **Cluster-corrected permutation p = 0.0020** |  |  |  |  |
| **UIUC** | 38 | 37 | −13.1 (−31.7 to 5.5) | 0.083 | −12.3 (−33.5 to 8.8) | 0.126 | 0.190 | −0.266 (−0.682 to 0.134) |
| **TCD** | 41 | 42 | −23.7 (−44.5 to −2.9) | 0.013 | −24.3 (−44.6 to −4.0) | 0.010 | 0.029 | −0.524 (−0.972 to −0.086) |
| **BCG** | 36 | 32 | −49.3 (−77.3 to −21.3) | <0.001 | −51.0 (−73.4 to −28.6) | <0.001 | <0.001 | −1.101 (−1.718 to −0.543) |
| **UZH** | 38 | 34 | −2.7 (−25.2 to 19.9) | 0.407 | −2.3 (−23.8 to 19.2) | 0.418 | 0.418 | −0.049 (−0.528 to 0.436) |
| **UKT** | 40 | 39 | −7.3 (−25.8 to 11.2) | 0.217 | −7.7 (−28.5 to 13.0) | 0.232 | 0.278 | −0.167 (−0.574 to 0.225) |
| **UR** | 39 | 39 | −22.4 (−41.2 to −3.6) | 0.010 | −20.8 (−41.8 to 0.1) | 0.025 | 0.051 | −0.450 (−0.871 to −0.051) |
| **All sites** | 232 | 223 | −19.4 (−28.1 to −10.8) | <0.001 | −19.3 (−28.2 to −10.4) | <0.001 | — | −0.413 (−0.607 to −0.233) |
| **— Site influence: site omission sensitivity —** |  |  |  |  |  |  |  |  |
| **omit UIUC** | 194 | 186 | −20.7 (−30.4 to −11.0) | <0.001 | −20.1 (−30.0 to −10.1) | <0.001 | <0.001 | −0.420 (−0.628 to −0.228) |
| **omit TCD** | 191 | 181 | −18.7 (−28.2 to −9.2) | <0.001 | −18.1 (−27.9 to −8.3) | <0.001 | <0.001 | −0.389 (−0.602 to −0.186) |
| **omit BCG** | 196 | 191 | −14.1 (−23.0 to −5.2) | <0.001 | −13.5 (−22.6 to −4.5) | 0.002 | 0.002 | −0.308 (−0.518 to −0.106) |
| **omit UZH** | 194 | 189 | −22.5 (−31.9 to −13.1) | <0.001 | −22.4 (−32.1 to −12.7) | <0.001 | <0.001 | −0.484 (−0.699 to −0.287) |
| **omit UKT** | 192 | 184 | −22.0 (−31.7 to −12.2) | <0.001 | −22.5 (−32.5 to −12.6) | <0.001 | <0.001 | −0.473 (−0.681 to −0.281) |
| **omit UR** | 193 | 184 | −18.7 (−28.4 to −9.0) | <0.001 | −18.8 (−28.8 to −8.8) | <0.001 | <0.001 | −0.394 (−0.602 to −0.190) |
| **— LOSO cross-validation: held-out site —** |  |  |  |  |  |  |  |  |
| **UIUC** | 38 | 37 | −13.1 (−32.7 to 6.5) | 0.093 | −11.7 (−31.8 to 8.4) | 0.125 | 0.188 | −0.268 (−0.688 to 0.193) |
| **TCD** | 41 | 41 | −19.8 (−39.0 to −0.6) | 0.022 | −19.0 (−38.2 to 0.1) | 0.026 | 0.154 | −0.437 (−0.869 to −0.017) |
| **BCG** | 32 | 30 | −27.4 (−66.7 to 11.8) | 0.084 | −27.1 (−66.4 to 12.2) | 0.087 | 0.173 | −0.350 (−0.860 to 0.137) |
| **UZH** | 38 | 34 | −7.7 (−28.9 to 13.4) | 0.234 | −7.5 (−28.7 to 13.8) | 0.243 | 0.243 | −0.165 (−0.637 to 0.302) |
| **UKT** | 40 | 39 | −7.4 (−27.7 to 12.8) | 0.233 | −8.8 (−29.3 to 11.7) | 0.197 | 0.237 | −0.193 (−0.617 to 0.253) |
| **UR** | 38 | 39 | −19.1 (−40.6 to 2.4) | 0.040 | −17.2 (−38.7 to 4.2) | 0.057 | 0.171 | −0.364 (−1.014 to 0.083) |
| **— Five-fold cross-validation: held-out fold —** |  |  |  |  |  |  |  |  |
| **Fold 1** | 47 | 44 | −15.6 (−35.8 to 4.6) | 0.064 | −14.1 (−34.0 to 5.7) | 0.080 | 0.101 | −0.297 (−0.668 to 0.063) |
| **Fold 2** | 47 | 44 | −23.6 (−52.6 to 5.3) | 0.054 | −22.4 (−51.8 to 6.9) | 0.066 | 0.101 | −0.319 (−0.690 to 0.073) |
| **Fold 3** | 46 | 45 | −7.7 (−27.7 to 12.3) | 0.223 | −7.6 (−27.6 to 12.4) | 0.225 | 0.225 | −0.159 (−0.531 to 0.238) |
| **Fold 4** | 46 | 44 | −15.0 (−32.8 to 2.9) | 0.049 | −13.8 (−32.0 to 4.5) | 0.069 | 0.101 | −0.316 (−0.663 to 0.062) |
| **Fold 5** | 46 | 45 | −21.9 (−42.4 to −1.4) | 0.018 | −22.0 (−42.9 to −1.0) | 0.020 | 0.100 | −0.437 (−0.820 to −0.071) |

Δ mean is the unadjusted difference in inhibition ratio between tinnitus and control participants, in percentage points; negative values indicate reduced inhibition in tinnitus. β is the covariate-adjusted group coefficient from ordinary least squares (×100, percentage points), adjusted for Age, Gender and PTA<sub>4</sub> within each site and additionally for Site in the pooled rows. Adjusted d is β divided by the model residual root-mean-square error; it is not a pooled-standard-deviation Cohen's d. All p values are one-sided in the pre-specified direction (tinnitus < control); confidence intervals are two-sided 95%, so an interval may include zero where p < 0.05. FDR q values are Benjamini–Hochberg adjusted p values computed separately within each block of this table; the pooled row is not a member of the per-site family and carries no q. The cluster-corrected permutation p for the pooled cluster-based contrast is given in the block header. Cross-validation rows re-derive cluster selection and the within-group outlier fence from training data alone, so a held-out participant contributes no score either because no electrode in the training-selected cluster meets the amplitude criterion or because their inhibition ratio falls outside the training-derived fence. These removed 1 and 7 participants respectively under leave-one-site-out (447 analysed) and 1 and 0 under five-fold cross-validation (454 analysed), from 455 entering each block.

No significant cluster inhibition was found in 3 or 8 kHz NBN, likely due to the overlapping of PO and GP N100 responses in either group (**Figure 2c**). The pre-specified hypothesis was that inhibition would be reduced in tinnitus at narrowband carriers centered near the tinnitus percept and preserved with broadband noise. However, the observed pattern was the opposite whereby inhibition was reduced with the broadband carrier and did not differ between groups at either narrowband carrier. With 229 cases and 226 controls at the 3 kHz carrier and 232 cases and 223 controls at 8 kHz, an effect of d = 0.233 would have been detected with 80% power, so the narrowband null is unlikely to reflect insufficient sample size alone. These findings strongly support the notion that tinnitus correlates specifically with an impaired inhibition of cGPIAS in a BBN carrier.

### Primary analysis: sensitivity of cGPIAS to tinnitus diagnosis

Raw N100 inhibition ratio (IR) alone showed modest discrimination between tinnitus participants and controls in the full cohort, with an apparent AUC of 0.610 (95% CI 0.559– 0.660). Combining IR with Site, Age, Gender, and PTA_4_ gave an AUC of 0.672 (0.622– 0.720; **Figure 3g**). Adding the 8 kHz hearing threshold (both ears’ average), psychological measures (PSQ, HADS-A, and HADS-D) yielded an AUC of 0.729 (0.682–0.773; **Figure S3a**). Pooled LOSO-CV and five-fold CV results for all models are reported in **Table S6.** Confidence intervals were estimated by conditional bootstrapping of the saved participant scores.

### Exploratory analyses: decreased inhibition in the matched cohort

As an exploratory analysis, we repeated the analyses in 199 tinnitus participants and 199 controls matched by gender, within five years of age and 5 dB PTA_4_. Cluster-permutation testing identified a similar 11-electrode frontocentral cluster, with FC6 replacing FC1 relative to the full-cohort cluster (cluster mass = 32.44; FWER-corrected permutation p = 0.0012; Figure **S4a,b**). The descriptive adjusted group difference was −24.61 percentage points (d = −0.552; Figure **S4c**), consistent with reduced inhibition in tinnitus in the full cohort. After omitting each site in turn, estimates within the fixed matched-cohort cluster remained negative (β range, −27.34 to −19.23 percentage points; d range, −0.615 to −0.468; all q < 0.001; **Figure S4d**).

In the matched cohort, LOSO-CV estimates were negative at all six held-out sites (β range, −33.86 to −16.60 percentage points; d range, −0.551 to −0.338), although FDR-adjusted q-values remained above 0.05 (q range, 0.0522–0.0931; **Figure S4e**). five-fold CV estimates were consistently negative (β range, −31.71 to −17.87 percentage points; d range, −0.499 to −0.357), with four of five folds surviving FDR correction (q = 0.0335) and the fifth falling just above the threshold (q = 0.0571; **Figure S4f**; **Table S7**).

Raw N100 inhibition ratio (IR) alone showed modest discrimination between tinnitus participants and controls in the matched cohort, with an apparent AUC of 0.642 (95% CI 0.587–0.696). Combining IR with Site, Age, Gender, and PTA_4_ gave an AUC of 0.663 (0.608–0.714; **Figure S4g**). Adding the 8 kHz hearing threshold and psychological measures yielded an AUC of 0.741 (0.693–0.788; **Figure S3b**). Pooled LOSO-CV and five-fold CV results for all models are reported in **Table S6.**

### Exploratory analyses: tinnitus severity correlates with the degree of impaired inhibition

The significant effect observed in BCG prompted us to examine in more detail the tinnitus variables across the different sites. Indeed, BCG is a tertiary clinic with cases whose tinnitus severity was greater than in any other site (see a companion paper by Shabestari et al.), suggesting that the impaired inhibition is related to severity. Among the 232 tinnitus participants analysed for the broadband carrier, THI scores fell in the negligible (0–16) band for 84 (36.2%), mild (18–36) for 92 (39.7%), moderate (38–56) for 45 (19.4%), severe (58– 76) for 9 (3.9%) and catastrophic (78–100) for 2 (0.9%). Among all eligible tinnitus participants, higher square-root THI (√THI) scores were associated with lower N100 inhibition ratios within the fixed full cohort 11-electrode cluster after adjustment for Site, Age, Gender and PTA_4_ (N = 213; β = −7.26 percentage points per unit √THI, 95% CI −11.71 to −2.81; two-sided OLS p = 0.0015; partial R² = 0.049; **Figure S5a**). This association persisted among participants with THI ≥ 18, with severity accounting for a greater proportion of residual variance in inhibition ratios (N = 134; β = −13.99 percentage points per unit √THI, 95% CI −22.50 to −5.48; p = 0.0015; partial R² = 0.079; **Figure S5b**).

### Exploratory analyses: reduced inhibition in tinnitus occurs in right temporoparietal regions

A covariate-adjusted vertex-wise Freedman–Lane Threshold-Free Cluster Enhancement (TFCE) analysis identified 2,064 significant vertices in the full cohort, including 1,835 in the right hemisphere and 229 in the left hemisphere (permutation-corrected p< 0.05; minimum corrected vertex p= 0.0022). All significant vertices showed lower inhibition ratios in tinnitus participants than controls. When focusing on clusters containing more than 100 significant vertices, two clusters comprising 1,616 and 165 vertices, both located entirely in the right hemisphere were found (**Figure 4**). The signal was centered on the right precuneus and extended through the superior parietal, middle and inferior temporal, fusiform, the banks of the superior temporal sulcus (bankssts), superior temporal, insular, and transverse temporal regions, indicating a continuous temporoparietal–auditory cortical distribution (**Figure 4b**).

**Figure 4.**
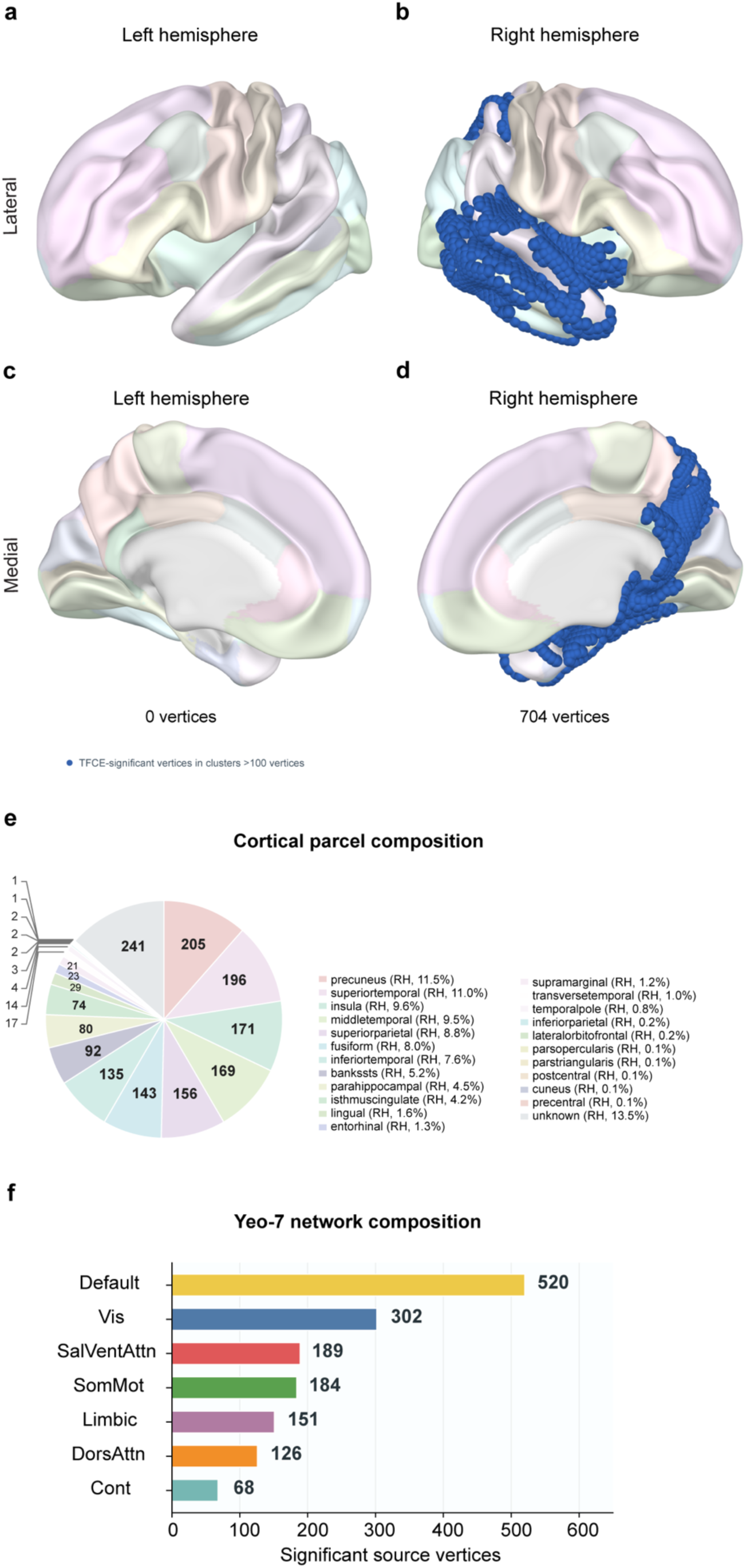
Whole-brain cortical distribution of significant vertices and Yeo-7 networks composition. Lateral (a, b) and medial (c, d) views of the left and right cortical hemispheres are shown. Translucent colored areas show Desikan–Killiany anatomical parcels. Blue dots indicate exact source vertices retained from TFCE-significant connected clusters containing more than 100 vertices. The retained clusters are confined to the right hemisphere, while the left-hemisphere panels show no retained significant vertices. The medial panels display the medial cortical surface and should not be interpreted as subcortical structures. (e) Number of retained TFCE-significant source vertices assigned to each Schaefer-100/Yeo-7 network. Bars are ordered from largest to smallest count. The Default network contains the most assigned vertices (520), followed by Visual (302), Salience/Ventral Attention (189), Somatomotor (184), Limbic (151), Dorsal Attention (126), and Control (68) and 241 vertices were unassigned to the seven networks. (f) Pie chart showing the cortical-parcel composition. Detailed parcel level counts and proportions are provided in **Table S8**.

The right precuneus and superior temporal cortex were the most prominent named anatomical territories, containing 205 and 196 significant vertices across the two clusters, respectively (**Table S8**). The peak TFCE vertex was located in the right precuneus (t = −4.62; vertex-wise FWER-corrected p = 0.0022), while the broader distribution also extended through lateral temporal and auditory cortical regions. Across the two large clusters, the assigned Yeo seven-network composition was Default (520 vertices), Visual (302), Salience/Ventral Attention (189), Somatomotor (184), Limbic (151), Dorsal Attention (126), Control (68) and 241 vertices were unassigned to a Yeo network (**Figure 4e,f, Table S8)**. This spatially organized pattern suggests that reduced inhibition in tinnitus is concentrated in right temporal and parietal regions rather than being diffusely distributed across the cortex.

## DISCUSSION

In a sample of 455 participants with and without tinnitus, we investigated whether the presence of subjective chronic tinnitus was associated with poorer inhibition of gap-mediated suppression of cortical sound-evoked responses from a diagnostic perspective. Patients with chronic tinnitus displayed significantly reduced inhibition of the N100 sound-evoked response when preceded by a gap in a BBN carrier in comparison to healthy controls. This difference was highly significant on a group level, but there was a high interindividual variability in the results in both groups, which severely limits its usefulness as an individual diagnostic marker.

Our source-localized deficit was confined to a predominantly right-hemisphere temporoparietal-auditory network, converging with prior evidence that right (but not left) secondary/associative auditory cortex is the physiological substrate of gap-mediated cortical inhibition. The involvement of the banks of the superior temporal sulcus (bankssts), superior temporal, and insular cortex suggests a broader distribution of reduced inhibition across auditory-related temporal and adjacent cortical regions rather than an isolated primary-cortex abnormality. Deeper structures were also identified. The prominence of right precuneus, the peak of our cluster, aligns with converging reports that tinnitus disrupts precuneus coupling to the default-mode and dorsal-attention networks, biasing patients toward a chronically "attentive" rather than resting cortical state^34^. Interestingly, the findings implicating precuneus were previously reported in structural MRI and resting-state fMRI meta-analyses^35^. This engagement in cGPIAS speaks to the role this region and the default mode network may play in the conscious awareness of tinnitus.

Parahippocampal involvement further implicates a memory-gating failure, whereby the phantom (or here, the un-gated pulse) response is not appropriately habituated or filtered as it would be in a healthy hippocampal-auditory memory loop^36,37^. Together, the default, dorsal attention, salience, limbic, and somatomotor network contributions in this cluster suggest that reduced inhibition in tinnitus is not restricted to auditory cortex but reflects a distributed excitation-inhibition imbalance spanning attentional, salience, and memory hubs, consistent with corticofugal and thalamocortical disinhibition models of tinnitus^38,39^. These findings extend prior GABAergic and connectivity work by directly linking impaired cGPIAS to this broader right-lateralized network architecture.

In animal models, behavioral GPIAS deficits have been reported in one or multiple high-frequency NBN conditions but seldom in BBN, the latter most often serving as a control for proper temporal processing^40^. Our protocol followed this reasoning, anticipating impaired gap inhibition at carriers centered near the tinnitus percept and no group difference with broadband noise, but the reverse was found. Inhibition was almost absent using NBN carriers in both groups. We believe that the lower amplitude of the gap-mediated ERP led to less efficient inhibition, plausibly because a 1 kHz-wide NBN excites a smaller extent of the tonotopic map than BBN. Consistent with this idea, manganese-enhanced MRI of the mouse brain shows that the volume of sound-activated tissue in the inferior colliculus scales with stimulus bandwidth, from broadband noise through high-frequency narrowband noise to a pure tone^41^. With so little inhibition to begin with, the dynamic range remained too small for a group difference to emerge. In addition, the fixed NBN center frequencies of 3 and 8 kHz may not have corresponded to individual tinnitus pitches, potentially obscuring frequency-specific effects. Differences in tinnitus loudness and frequency-specific hearing thresholds may have further contributed to variability in cortical responses. The prediction of impaired inhibition at individually tinnitus-matched frequencies therefore remains untested rather than refuted. The consistency of the direction of the effects across all six contributing sites, recorded on different equipment and by different teams, argues against a site-specific or equipment-specific artefact. Whether the effect survives prospective testing remains to be determined in a study in which broadband is the a priori carrier and the region of interest is fixed in advance.

A generalized reduction of cortical inhibition in tinnitus, affecting both pre- and post-gap activity across a broad tonotopic region could be expected to alter the relative salience of the post-gap responses for any stimulus that strongly activates auditory cortex, including a BBN^42^. Consequently, we speculate that our BBN findings reflect a greater sensitivity of the human auditory cortex to a more tonotopically-spread disinhibition of this circuit. Consistent with a recent study by Morse et al.^43^, BBN may be sufficient to assess global auditory network excitability and gating to detect an aberrant state of the whole auditory network as occurring in individuals with tinnitus. This would considerably decrease the assessment duration and allow for MEG or OPM-MEG measures that are far more sensitive than EEG to capture impacted cortical inhibition^44^.

It has also been suggested that the tinnitus percept “fills in” the silent gap, thereby impairing the proper inhibition of the startle response. However, both animal and human studies indicate that this latter mechanism may not explain reduced GPIAS as both species still perceive a gap even when presented with a carrier noise including a frequency closely matching that of the putative tinnitus pitch^25,45^. In contrast, our study strongly supports the notion that a reflex mechanism, cortically mediated and acting far earlier than the conscious percept, underlies the GPIAS response. Importantly, while our results are not a direct validation of the numerous animal experiments that were performed using GPIAS and corroborated against neural correlates of tinnitus, they are in strong support. GPIAS in animals remains highly debated and poorly trusted^40^, and the equivalent blinking response in humans cannot be measured in nearly half of subjects, even with high stimuli^33^. In contrast, previous pilot MEG studies and the present study support cortical measures as an alternative to the blinking response, which could be back translated in animals^46^ to be refined into a more clinically valid, objective measure of tinnitus. Several clinical trials on tinnitus using direct vagal nerve stimulation^47^, or bimodal stimulation^11^, relied on animal experiments using GPIAS^10,23^. Should these interventions be validated using cGPIAS, the proposed methodology may be used for drug discovery and interventional trials.

Can the cGPIAS paradigm be used as a diagnostic marker? Likely not. The observed variability suggests it would be insufficiently sensitive for diagnosing an individual with tinnitus. Within TIDE, the two companion paradigms set a useful benchmark. Under the same leave-one-site-out scheme, covariate-adjusted cGPIAS inhibition reached a held-out AUC of 0.585, and 0.616 in the age- and hearing-matched cohort, whereas the multimodal resting-state normative model reached 0.819, rising to 0.832 for the soft-voting ensemble (companion paper by Shabestari et al.), and the auditory oddball paradigm reached 0.665 (companion paper by Shields et al.). The comparison is not like-for-like, since the resting-state estimate derives from a 680-feature multimodal ensemble while cGPIAS rests on a single cortical measure. Nonetheless, spontaneous oscillatory deviation carries more information about the presence of tinnitus than the gap-evoked cortical response does. However, the dissociation reverses for severity. In the resting-state cohort, regression of THI and TFI from the same features returned negative cross-validated R^2^, and the strongest single-feature association with distress did not survive correction, so that signature indexes the percept rather than its burden. cGPIAS behaves in the opposite way, scaling with handicap within cases (partial R^2^ = 0.049 across all eligible tinnitus participants, and 0.079 restricted to those with THI ≥ 18). Detection and severity monitoring may therefore demand different measurements, and an evoked response that varies with the degree of handicap is the more plausible candidate for tracking change within an individual over time, which is what a trial outcome measure requires.

Accordingly, cGPIAS may serve as a surrogate measure of severity at a group level, replacing the subjective measure of tinnitus distress (e.g., THI) used in nearly half of tinnitus Randomized Controlled Trials (RCTs). GPIAS studies in mice are also in support of this hypothesis. Young adult mice lacking the glutamate aspartate transporter (GLAST) show normal GPIAS and normal behavioral hearing thresholds at baseline, despite displaying auditory synaptopathy in the absence of any noise exposure with halved auditory synapses and reduced ABR Wave 1 amplitude when compared to wild-type mice^48^. Only when challenged with high doses of salicylate causing temporary tinnitus, was GPIAS reduced by 80% in knock-out mice in comparison to 25% in wild-type mice, suggesting that salicylate caused mild tinnitus and that synaptopathy exacerbated this phenotype. While these results in GLAST KO mice indicate that auditory synaptopathy is necessary but not sufficient for tinnitus development, they also suggest that a weakened cochlear output synchrony of high-spontaneous firing rate fibers could impact cortical inhibition processes^49,50^, providing a likely explanation of the observed changes in cGPIAS in humans with tinnitus from the present study.

We acknowledge both strengths and limitations of this work. The analyzed sample comprises 455 participants from academic, medical and tertiary clinics from six sites across two continents (e.g., tinnitus characteristics, demographics, and distress patterns differed between sites), collected by multiple staff (e.g., medical nurses, medical students, master students), and recorded on different EEG devices. The electrode cluster used for the primary analysis was identified from the same contrast in which it was subsequently tested, so the effect size at that cluster is optimistically biased. In contrast, the spatio-temporal analysis, which was data-driven across the full montage, provides the less circular estimate. Analyses across carriers, electrode clusters, sites, folds and source vertices were not corrected as a single family; cluster-level and false-discovery-rate corrections were applied within each analysis, and the exploratory results should therefore be read as hypothesis-generating. The direction of the effects was found consistent across sites, even in those with greater tinnitus distress (and thus more clinically relevant), yielding strong aggregated evidence with an effect size that increases with tinnitus severity. Unfortunately, our sample was poorly represented with severe tinnitus patients. This could be due to the fact we excluded several participants with hyperacusis for safety reasons. Because hyperacusis is tightly associated with severe tinnitus^51^, it is possible that our exclusion criteria impacted the recruitment of participants that would have otherwise had clinically relevant tinnitus. On the positive side, none of the participants reported the paradigm to be unpleasant. Although not consistently recorded, numerous patients that sought contact within the clinics to be enrolled in the study were not included due to severe hearing loss from their medical records. In future interventional trials, it will be critical to adjust the carrier level to allow the assessment of more hearing-impaired individuals. While this study could not reliably diagnose tinnitus at the individual level, it suggests a novel objective biomarker at the group level for chronic tinnitus in humans. The present internal and external validity provides a rationale to conduct prospective interventional trial on diverse tinnitus populations. Further research is needed to optimize the sensitivity of the paradigm, potentially with MEG or OPM-MEG and determine its potential use as a trial outcome measure for tinnitus intrusiveness.

## METHODS

The goal of TIDE is to investigate, in a large international sample of tinnitus patients and controls, the sensitivity of various paradigms in diagnosing tinnitus in humans, namely the oddball paradigm and cortical GPIAS^33,52^. The paradigms were modelled in TIDE using free software for analysis (Python), available codes in repositories (GitHub) and various EEG hardware for a facilitated implementation in diverse clinical settings. The present study only analyzed the data collected on the GPIAS paradigm. Study oversight was conducted by a single institution (Trinity College Dublin). We followed the Standards for Reporting of Diagnostic Accuracy Studies (STARD) and the Strengthening the Reporting of Observational Studies in Epidemiology (STROBE) reporting guidelines^53–55^. Reporting guidance for multivariable prediction models was not applied, because the index test is reported as a single covariate-adjusted measurement rather than as a model combining predictors to yield individual probabilities.

### Study Planning and Conduct

TIDE is a prospective, multicentre case–control study conducted at seven clinical sites in the United States and Europe between September 2024 and October 2025. Five European sites were identified on the basis of expertise and track record in tinnitus research. The two US sites were identified by the European investigators, to enable future implementation under NIH guidelines. The participating sites were the University of Texas at Austin (UTA, USA), University of Illinois Urbana-Champaign (UIUC, USA), Trinity College Dublin (TCD, Ireland), Brai3n Clinic (BCG, Belgium), University Hospital Zurich (UZH, Switzerland), University Clinic Tübingen (UKT, Germany), and the Tinnitus Clinic at University Hospital Regensburg (UR, Germany). The protocol is published^56^.

### Ethics, registration and study size

The study has been approved by the respective Ethics committees or Institutional Review Boards at the seven sites (UTA: #STUDY00004651; UIUC: #IRB24-1117; TCD: # SPREC022020-07; BCG: # 6497-B3002024000114; UZH: #KEK-ZH 2025-01839; UKT: #079/2024BO2; UR: #23-3580-101) and is compliant with US and European data protection regulations. Written informed consent was obtained from every participant before any study procedure, in accordance with the Declaration of Helsinki. The study has been pre-registered on ClinicalTrials.gov (NCT06520865). The study was registered before enrolment, but the protocol did not designate primary or secondary outcome measures or a statistical analysis plan. The group contrast in each carrier and the diagnostic accuracy of the broadband contrast are designated here as the primary analyses. All other analyses are exploratory and are reported as hypothesis-generating. No a priori power calculation was performed. No prior estimate of the effect of tinnitus on cortical gap-induced inhibition was available, this being the first study to measure GPIAS through cortical evoked responses at this scale, so no effect size could be assumed. The target of 40 cases and 40 controls per site was set by the enrolment achievable across the seven sites during the recruitment window. The power statement in the published protocol was included in error, and a correction has been requested. In the analyzed sample of 232 cases and 223 controls, an effect of d = 0.233 would be detected with 80% power at a one-sided α of 0.05.

### Participants

Participants were recruited to tertiary or academic tinnitus clinics and research centers via institutional emails, flyers, and journal announcements (**Figure 1c**). Participants therefore formed a convenience series rather than a consecutive or random sample of the clinic populations. Upon arrival at the clinical centers, participants received detailed information about the study procedures and provided written informed consent. Eligible participants were aged 18 to 70 years, able to understand and consent to the research, cognitively unimpaired (Montreal Cognitive Assessment ≥ 26, presented only by certified researchers) and without hypersensitivity to sound (Hyperacusis Questionnaire ≤ 27; a score of 28 or above, the instrument’s own threshold for hyperacusis, was exclusionary). UZH administered a German version of the HQ questionnaire with three response options per item instead of four-option version used at the other six sites. As a consequence, no participant was excluded on this criterion at UZH. Exclusion criteria were objective tinnitus, heartbeat-synchronous tinnitus as primary complaint, otosclerosis, acoustic neuroma or other relevant ear disorders with fluctuating hearing, acute ear nose and throat infections, Meniere’s disease or similar syndromes, vestibular migraine, serious internal, neurological or psychiatric conditions, epilepsy or other central nervous system disorders, clinically relevant drug, medication or alcohol misuse within 12 weeks of study start, severe hearing loss, and single-sided deafness. After their inclusion, participants underwent an on-line survey and audiological assessments prior the EEG recording session. After completing all measurements, a financial compensation, adjusted to local costs of living, was provided.

The survey confirmed none had been diagnosed with Ménière’s Disease, acoustic neuroma, or otosclerosis. Less than 10% of the participants had acoustic trauma, acute or chronic otitis, or sudden hearing loss as a potential cause of their tinnitus. Eight and five participants reported chemotherapy and head and neck radiotherapy respectively (**Table S2**). Of 820 individuals assessed for eligibility, 236 were excluded and 584 were enrolled (**Figure 1c, Table S3**).

Reasons for exclusion by site are given in **Table S4**. A complete record across questionnaires, audiometry and EEG was available for 562 of the 584. The analyses reported here draw on this eligible sample on whom a quality control (QC) described below was applied. The final TIDE pool of analyzable clinical and electrophysiological cGPIAS data after QC comprised 233 chronic tinnitus cases and 224 non-tinnitus controls for a BBN carrier. Two participants (one tinnitus and one control) were subsequently excluded from the statistical analyses due to incomplete PTA_4_ data in one ear, resulting in a final analytical sample of 455 participants (232 tinnitus and 223 controls).

The protocol aimed to match cases and controls at group level for age, sex and hearing, however this was not achieved. Preserving the match would have required declining eligible participants during recruitment, which was not operationally feasible, and the intent was in any case difficult to reconcile with the well-established difference in hearing between people with and without tinnitus. Age, gender, PTA_4_ and site are therefore carried as covariates in all adjusted analyses rather than relied upon as balanced. Site differences in recruitment, questionnaire and audiometric profiles will be reported separately.

### Questionnaires and Audiometry

Included participants, both cases and controls, completed the European School for Interdisciplinary Tinnitus Survey Questionnaire (ESIT-SQ) Part A, Hospital Anxiety and Depression Scale (HADS), and the Perceived Stress Questionnaire (PSQ-30). For cases only, the ESIT-SQ Part B, the Tinnitus Handicap Inventory (THI), and the Tinnitus Functional Index (TFI) were used. Questionnaires were delivered either on paper or electronically via REDCap, with the exception of UZH, which collected survey data on their own platform UniPark. Data from UniPark was recoded for a merge with Redcap data. A summary of the original questionnaire source and their reliability (Cronbach’s α) when delivered electronically is available in Müller et al.^57^ THI distress grading was used to classify tinnitus according to negligible distress (0-16), light weight (18-36), moderate weight (38-56), severe weight (58-76), and catastrophic (78-100)^58,59^. There were fifteen missing entries on the PSQ score and controls from Zurich did not answer the ESIT-SQ (n = 36 missing).

Hearing thresholds were evaluated using pure-tone audiometry at 0.25, 0.5, 1, 2, 3, 4, 6, 8 and 12.5 kHz. PTA_4_ values were extracted from the average of the 0.5, 1, 2, 4 kHz thresholds. Whenever a device would allow so, extended high frequency (eHF) audiometry was collected from 10 up to 16 kHz. Audiometers differed between sites, as did the frequencies each could deliver and its maximum output at each frequency (**Table S5**). Thresholds at 8 and 12.5 kHz were obtained at every site, and their average is reported as the high-frequency average. At UKT the audiometer reached 16 kHz but was found to deliver a narrowband noise rather than a pure tone at that frequency. This frequency was therefore withdrawn at that site, and 14 kHz is the highest frequency for which UKT thresholds exist.

Where a participant did not respond at a site’s maximum output, the threshold was recorded as that maximum plus 5 dB, a convention agreed across the consortium and standard in clinical screening. Such corrections only occurred at 8 kHz and above, rising from 0.7% of ear measurements at 8 kHz to 29.0% at 16 kHz (**Table S5**). The high-frequency average was evaluated as an additional hearing covariate and did not materially alter the models. PTA_4_, which is unaffected, was retained as the hearing covariate. Tinnitus pitch and loudness matching were assessed using a paired-comparison procedure to determine the perceived tinnitus frequency and corresponding loudness. Sound intensity and frequency sensitivity for tinnitus psychoacoustics differed between the hardware used at each site (**Table S2)**.

### Reference standard

Tinnitus status, the target condition, was chronic, constant, subjective, non-pulsatile primary tinnitus. Case and control status was established at each site by clinical interview, conducted by an audiologist at the US sites and by a physician at the European sites. Cases required a diagnosis of chronic tinnitus of at least six months’ duration based on history, perceived as constant, subjective and non-pulsatile, and classified as primary tinnitus. Tinnitus characteristics were documented with ESIT-SQ Part B. Case and control status was established at the eligibility interview, before any EEG was recorded and within the same study visit, so index-test results were not available to the assessors of the reference standard and no intervention intervened between the reference standard and the index test. Controls required no experience of tinnitus lasting longer than two minutes on any single occasion.

This is the operationalization of the registered criterion, which excludes anyone who has experienced tinnitus other than short-term. No objective confirmatory test for tinnitus exists, so the reference standard is standardized self-report obtained at clinical interview. This is the limitation the TIDE program sets out to address, and it bounds the accuracy attainable by any index test, since misclassification at the reference standard propagates directly into estimated sensitivity, specificity and discrimination.

### Primary Outcome

The primary outcome was the difference in the inhibition ratio of the N100 mean amplitude in a BBN, 3 kHz, or 8 kHz NBN carrier between cases and controls.

#### EEG design, preparation and recordings

Before EEG recording, all participants were instructed to remove metal objects (e.g., hairpins, piercings, jewelry). Standard EEG preparation was performed, including scalp cleansing and placement of the EEG cap. EEG data were collected using 64-channel systems (**Figure 2a, Table S1**). To minimize sleepiness during exposure to the white noise carriers and distraction from other objects in the chamber, participants were instructed to sit comfortably and fixate on a cross displayed on a monitor in front of them. The EEG session began with a 5-minute resting-state recording in silence with eyes open. During the GPIAS paradigm, all auditory stimuli were presented binaurally through insert earphones.

### GPIAS paradigm

Sounds were calibrated at each site using similar protocols, although with different calibration devices, and delivered binaurally through ER2 (Etymotic) or EARtone GOLD 3A (3M) ear inserts with a background sound pressure level of <35 dB(A) sound pressure level (SPL), shortened as dB. Broadband sound pulses (20 ms, 90 dB, no rise and fall time) were presented in a continuous 60 dB carrier broadband noise (BBN), or a 1/3-octave bandpass-filtered narrowband noise (NBN) centred at 3 kHz or 8 kHz. This was done to closely match the analogue Butterworth filtering (48 dB/octave ramp) we previously used in animal studies^60^.

Silent gaps (50 ms, 2 ms rise and fall ramp, down to background noise floor level) preceded a pulse by 240 ms interstimulus interval (ISI). Trials were 3.064 s long containing either a pulse only (PO), a gap and a pulse (GP), or a gap only (GO) (**Figure 2b)**. GO trials served as an internal control to confirm the absence of pulse evoked responses after a gap ERP. The measures consisted in five sessions, with session 1 and 5 containing PO trials with pulses at 70, 75, 80, 85, 90 dB randomly presented 25 times each allowing for verifying habituation at the end of the assessment. Sessions 2, 3 and 4 contained blocks of BBN, 3 kHz or 8 kHz with PO, GP, and GO randomly presented 100 times each (**Figure 2c)**. The order of sessions 2, 3, 4 was also randomized. Following every trial in every session, a randomized inter-trial jitter (still containing the carrier sound) ranging from 1 to 500 ms was presented. The grand average for PO and GP trials across all sites is shown in **Figure 2d**. After each block, we communicated with the participant via an intercom system to make sure they were comfortable and assessed sleepiness. The total scanning time was around 60 min.

### EEG preprocessing

All EEG data were pre-processed using MNE-Python^61^. Recordings from all sites were mapped onto a common standard 64-channel montage (EasyCap M1). Continuous EEG was resampled to 250 Hz, re-referenced to the average of all electrodes, and band-pass filtered (1– 40 Hz). Noisy channels and residual artifactual segments were identified visually and removed from the continuous recording. Ocular artifacts were removed using independent component analysis (infomaxICA from *mne.preprocessing.ICA*) and flagged channels were interpolated. For the GPIAS paradigm, epochs were extracted from −200 to 500 ms relative to trigger onset, and epochs with residual artifact were excluded following automated and visual review. Evoked responses were computed by averaging artifact-free epochs within each condition. No baseline correction was done here because it would have overlapped with gap-evoked responses. The analyst was blinded to group.

### Cortical GPIAS quality control

Site-specific grand averages and topographies were inspected for morphology, latency, and signal-to-noise ratio of N100 responses for each carrier and condition. Qualitative investigation of the ERPs indicated that UTA failed in capturing responses to pulses in GP trial, whatever the carrier being presented. A retrospective investigation of the anomalous EEG recordings at UTA (e.g., inspection of raw data, event logs, and acquisition hardware) revealed a failure of the trigger circuitry. To avoid biasing the aggregated analyses, we therefore excluded UTA from all EEG analyses.

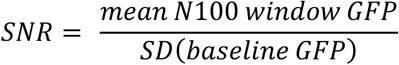

To ensure sufficient signal quality for the quantification of N100 amplitude and its inhibition ratio, ERPs were assessed for each participant and per carrier condition (BBN, 3 kHz, 8 kHz) using global field power (GFP) followed by the root-mean-square amplitude across all EEG channels ^62,63^, computed on the PO trials evoked response. This channel-agnostic measure was chosen over a spatial region of interest to avoid circularity with the topographic analyses described below. Signal-to-noise ratio was defined as mean GFP in the N100 window (60– 140 ms post-stimulus) divided by the standard deviation of GFP in the pre-stimulus baseline (−200 to 0 ms). Participants with an SNR ≥ 2.0 per carrier were kept for further analyses. This SNR cut-off was not pre-specified and was deducted from the data. Recordings failing this criterion were treated as indeterminate index-test results; they were neither reclassified nor imputed and contributed to no analysis. Testing an SNR ≥ 1.5 yielded similar outcomes, however, we chose the most conservative high-quality data approach. After quality control (QC), 457 participants passed the SNR criterion for each carrier: 233 tinnitus and 224 control for BBN, 230 tinnitus and 227 control for 3 kHz NBN, and 233 tinnitus and 224 control for 8 kHz NBN. Quality control was applied separately to each carrier, so these three samples differ in composition: 449 participants passed all three carriers and 461 passed at least one. In two participants, audiometric thresholds had not been recorded in one ear, so no binaural four-frequency pure-tone average could be derived; both were therefore removed by listwise deletion from every covariate-adjusted model, giving analyzed samples of 455 for BBN (232 tinnitus, 223 control), 455 for 3 kHz NBN (229 tinnitus, 226 control) and 455 for 8 kHz NBN (232 tinnitus, 223 control).

### Statistical Analysis

#### Primary outcome

For each cluster identified above, we used the shared grand-average N100 peak latency to compute a per-subject scalar inhibition ratio from the peak-window mean amplitude (± 20 ms). Observations required PO amplitude >0.1 µV and outliers were identified using a 3×IQR Tukey fence computed separately within the tinnitus and control groups and excluded from the regression. In the cross-validation analyses this fence was derived from training data only and applied unchanged to held-out participants. All models were fitted on complete cases and no values were imputed^64,65^. Group differences were estimated by covariate-adjusted OLS:

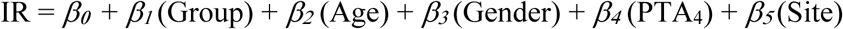

Age and PTA_4_ were each entered either as a linear term or as a natural cubic spline, with the functional form chosen by AIC. AIC selected the linear form for both, so the model reported here is linear in age and PTA_4_.

### Spatial cluster-based permutation analysis

Spatial cluster-based analyses were performed separately for each carrier condition (BBN, 3 kHz and 8 kHz). Peripheral electrodes prone to myogenic artifacts were excluded, and analyses used the retained 41-electrode montage. N100 peaks were identified separately for PO and GP at each electrode from group-pooled grand-average waveforms within 60–140 ms. Mean amplitudes within ± 20 ms were converted to absolute values before calculating inhibition ratio. Group differences were tested using OLS Freedman–Lane residual permutation testing, with Gender, Site, Age and PTA_4_ included in the regression model.

Residuals from the reduced model excluding Group were permuted among eligible participants at each electrode. Negative clusters required one-sided p <0.05 and at least two neighboring electrodes. Cluster mass was calculated by summing negative t-statistic magnitudes, and cluster-corrected p-values were obtained using 5,000 permutations and the maximum cluster mass across all 41 electrodes. Because the electrode cluster was identified from the same contrast in which the group difference was subsequently estimated, effect sizes at that cluster are optimistically biased; the cross-validation analyses described below re-derive cluster selection within training samples and provide the less circular estimate.

LOSO-CV and five-fold CV analyses repeated window estimation, filtering and cluster selection within training samples^66,67^. Training-derived clusters and nuisance adjustments were applied to held-out participants, and group differences were evaluated using one-sided OLS regression tests. Benjamini–Hochberg FDR adjustment was applied separately within each family of site-specific, LOSO-CV and five-fold CV comparisons.

#### Primary analysis: sensitivity of cGPIAS to tinnitus diagnosis

ROC (Receiver Operating Characteristic) curves were constructed from participant-level mean inhibition ratios across eligible electrodes in the conventional OLS Freedman–Lane-selected clusters. IR scores were adjusted for gender, site, age and PTA_4_. Group differences were calculated as tinnitus minus control, with negative values indicating lower inhibition in the tinnitus group (one-sided).

Discrimination was summarized by the area under the ROC curve (AUC). No positivity threshold was defined: the index test is reported as a continuous covariate-adjusted score, and no classification of individual participants was performed. LOSO-CV and five-fold CV ROC analyses were performed separately in the full cohort and the matched cohort. Apparent AUCs used cluster selection and adjustment estimated within that entire cohort. For LOSO-CV and five-fold CV analyses, N100 windows, cluster selection and adjustment were estimated using training participants and applied unchanged to held-out participants. Held-out scores were then pooled to calculate AUC separately for each cohort and validation design. Conditional 95% confidence intervals were estimated using 20,000 bootstrap resamples, preserving site and group counts in the full cohort.

*Exploratory analyses: matched cohort.* The analyses were repeated in 199 tinnitus–control pairs matched within five years of age and 5 dB PTA_4_, allowing cross-site matches. The same measurement, filtering, covariate-adjustment, and statistical procedures were applied, with cluster selection repeated within the matched cohort. Site-influence analyses retained the matched-cohort cluster while excluding each site in turn. LOSO-CV used entire sites as held-out samples, whereas five-fold CV kept matched pairs within the same fold. FDR adjustment was performed separately within each matched-cohort comparison family. Conditional 95% confidence intervals for AUC were estimated using 20,000 percentile-bootstrap resamples of matched pairs.

#### Exploratory analyses: Continuous regression analysis of tinnitus severity

Among tinnitus participants, the association between square-root THI (√THI) scores and inhibition ratios within the fixed full-cohort 11-electrode cluster was examined using OLS regression adjusted for site, gender, age and PTA_4_. Analyses included all eligible tinnitus participants and were repeated among those with THI ≥18, retaining the existing eligibility and outlier criteria. THI is a bounded sum of 25 ordinal items with a floor at zero and was right-skewed among the 232 tinnitus participants analyzed for the broadband carrier (skewness +0.81; Shapiro–Wilk W = 0.95, p < 0.001), so the untransformed score places disproportionate leverage on the few participants with severe handicap. The square-root transformation removed this skew (skewness −0.08; W = 0.99, p = 0.54) and reduced the share of predictor variance carried by the five most extreme observations from 19.1% to 13.8%, whereas a logarithmic transformation over-corrected (skewness −1.01). The transformation was chosen on these distributional grounds and was not pre-specified. Coefficients were reported per unit √THI, with conventional OLS 95% confidence intervals, two-sided p values and partial R2.

### Exploratory analyses: Source reconstruction

Cortical source activity was estimated on the FreeSurfer fsaverage template^68^. The forward model was generated using a three-compartment boundary element model representing the scalp, outer skull, and inner skull surfaces (*mne.make_forward_solution*). Sensor noise covariance was estimated separately for each participant from the pre-stimulus periods of the single-trial EEG epochs (*mne.compute_covariance*). Participant-specific minimum-norm inverse operators were subsequently constructed (*mne.minimum_norm.make_inverse_operator*), and source estimates were obtained using dSPM noise normalization (*mne.minimum_norm.apply_inverse*).

For the present source-level analysis, cortical source estimates were analyzed at the source-vertex level within the 60–140 ms N100 search window. For each participant and source vertex, the inhibition ratio was computed as (IR = 1 - |GP|/|PO|) from the PO and GP source amplitudes. The previously described outlier-removal and covariate-adjustment procedures were applied consistently. Vertex-wise group differences were tested in the one-sided tinnitus < control direction using Freedman–Lane residual permutation testing with TFCE and 5,000 permutations and significant vertices were defined as vertex-wise FWER-corrected TFCE p < 0.05^69,70^. Spatial clusters were defined descriptively as connected sets of significant vertices. Clusters containing more than 100 vertices were presented for anatomical visualization and summary. Desikan–Killiany labels described anatomical composition^71^, while Schaefer-100/Yeo-seven-network labels described network composition^72,73^. These summaries did not constitute separate parcel-level or network-level significance tests.

## Data availability

Anonymized individual participant data (questionnaire, audiometry, EEG) may be made available on reasonable request to the corresponding author, subject to institutional approval and a data sharing agreement. A consortium-wide release is planned once the combined machine-learning analysis of all TIDE paradigms is complete; no date is yet fixed.

## Code availability

The codes used for presenting the paradigm, analyzing EEG data and generating figures, are available in the following repository (https://github.com/orgs/TIDEConsortium/).

## Contributions

Conceptualization: T.K., F.T.H., J.C., D.D.R., B.L., W.S., S.V., P.K.A.N., C.R.C. Data Curation: H.L., P.S.S. Formal Analysis: H.L., P.S.S. Funding Acquisition: T.K., F.T.H., J.C., D.D.R., B.L., W.S., S.V., C.R.C. Investigation: H.L., M.E., S.A., E.A., E.K., F.C., N.J., J.O., J.K., S. R., L.R.R., I.S.F.C., N.S., S.M.W., A.Y.-M. Methodology: H.L., P.S.S., P.K.A.N., C.R.C. Project Administration: T.K., S.S., S.M.W., F.T.H., J.C., D.D.R., B.L., W.S., S.V., C.R.C. Resources: H.L., C.B., T.K., S.M.W., F.T.H., J.C., D.D.R., B.L., S.V., P.K.A.N., C.R.C. Software: H.L., P.S.S., P.K.A.N. Supervision: T.K., S.M.W., F.T.H., J.C., D.D.R., B.L., S.V., P.K.A.N., C.R.C. Validation: H.L., M.E., S.S. Visualization: H.L., P.S.S., P.K.A.N., C.R.C. Writing – Original Draft: H.L., C.R.C. Writing – Review & Editing: H.L., P.S.S., M.E., S.A., E.A., C.B., F.C., N.J., E.K., T.K., J.K., L.R.R., I.S.F.C., N.S., S.S., B.T., S.M.W., A.Y.-M., F.T.H., J.C., D.D.R., B.L., W.S., S.V., P.K.A.N., C.R.C. All authors played a role in writing the manuscript and approved the final version.

## Acknowledgements

We thank all the participants who joined the study. We also thank Marlies Knipper, Lukas Rüttiger, Stefan Fink and Rodrigo Andrès Donoso-San Martin for their support in the recruitment procedures at UKT, as well as Aurore Brugeaud and Karina Gültig for their administrative support. This project was supported by the Rainwater Charitable Foundation (#18340) and the American Tinnitus Association. C.R.C. received additional funding from the Swedish Research Council (Vetenskapsrådet #2023-02326). P.S.S was also supported by the Swiss National Science Foundation (SNF Project Grant number 208164).

## Competing interests

C.R.C. is member of the Professional Advisors’ Committee from the Tinnitus UK and member of the Scientific Advisory Board of the American Tinnitus Association, Swiss Tinnitus Liga, and Tinnitus Quest. C.R.C. is Editor-in-Chief of the *Journal of the Association for Research in Otolaryngology* (JARO) and receives a stipend from the Association for Research in Otolaryngology (ARO). F.T.H. is a member of the Scientific Advisory Board of Hyperacusis Research and SoQuiet Foundation. J.C. is a member of the Scientific Advisory Board for the Misophonia Research Foundation and a member of the Scientific Advisory Board of the American Tinnitus Association. B.L. received research support from the European Union, Neuromod and Sonova, honorary for consultancy and lectures from Neuromod, Schwabe, Sonormed, Sea Pharma, Sonova and consumer protection organizations.

## Role of the Funder/Sponsor

The Rainwater Charitable Foundation initiated the project and identified the investigators who formed the consortium. The study design and methods were developed by the academic investigators. Representatives of the Foundation attended consortium meetings. The Foundation had no role in the collection, management, analysis or interpretation of the data, in the preparation, review or approval of the manuscript, or in the decision to submit the manuscript for publication.

## Notes

### Clinical Trial

NCT06520865

### Clinical Protocols

https://bmjopen.bmj.com/content/16/5/e112788

### Author Declarations

The study has been approved by the respective Ethics committees or Institutional Review Boards at the seven sites (UTA: #STUDY00004651; UIUC: #IRB24-1117; TCD: # SPREC022020-07; BCG: # 6497- B3002024000114; UZH: #KEK-ZH 2025-01839; UKT: #079/2024BO2; UR: #23-3580-101) and is compliant with US and European data protection regulations. Written informed consent was obtained from every participant before any study procedure, in accordance with the Declaration of Helsinki. The study has been pre-registered on ClinicalTrials.gov (NCT06520865).

### Summary of Updates

Receiver operating characteristic analysis. The previous version reported a single covariate-adjusted discrimination estimate for the broadband inhibition ratio (apparent AUC 0.612, 95% CI 0.560 to 0.663). This version reports the unadjusted inhibition ratio as the primary index (apparent AUC 0.610, 95% CI 0.559 to 0.660) and adds a model ladder: adding site, age, gender and the four-frequency pure-tone average gives 0.672 (0.622 to 0.720), and further adding the 8 kHz hearing threshold and psychological measures (PSQ, HADS-A, HADS-D) gives 0.729 (0.682 to 0.773). The corresponding matched-cohort values are 0.642, 0.663 and 0.741. Held-out leave-one-site-out and five-fold cross-validated AUCs for every model are now tabulated in a new Supplementary Table S6, and the added-variable ROC curves are shown in a new Supplementary Figure S3. The conclusion is unchanged: discrimination of individuals remains limited. Abstract. The reported AUC is now identified as unadjusted and updated to 0.610 (95% CI 0.559 to 0.660). Matched cohort. The matching criteria are now stated in full: pairs were matched by gender in addition to within five years of age and 5 dB of four-frequency pure-tone average. Figures and tables. Figure 3 and the corresponding matched-cohort supplementary figure have been regenerated on a common scale, and Supplementary Table S7 has been updated. Reporting checklists. The STROBE and STARD checklists in the Supplementary Materials have been updated to this version. All line references were recomputed against the present line-numbered PDF, the reported areas under the curve were corrected to match Supplementary Table S6, and the vertex-wise source reconstruction analysis, Figure 4 and Supplementary Table S8 were added to the items covering subgroup, sensitivity and variability analyses. Supplementary Materials. The contents page now carries page numbers. The contents entry for Supplementary Table S6 has been corrected, having previously duplicated the title of Table S7. Methods. The spatial cluster-based permutation analysis subsection has been repositioned, and minor wording and style corrections were made throughout. No results or conclusions are affected by these changes.

