## Supplementary material for "Impaired cortical gap-mediated inhibition in chronic tinnitus but with limited individual diagnostic accuracy"

*Study registration: NCT06520865*

---

**Table S1. Equipment and devices across the 7 TIDE sites**

| | | UTA | UIUC | TCD | BCG | UZH | UKT | UR |
|---|---|---|---|---|---|---|---|---|
| Audiometry | Headset | RadioEar DD450 | HDA200 | HDA300 | HDA200 | HDA200 | HDA300 | HDA300 |
| | Audiometer | GSI (Audiostar) | Equinox (Interacoustics) | Hekto (Horentek) | (MedRx) | RME UCX II | AT1000 (Auritec) | Astera 2 (Otometrics) |
| | dB steps (dB) | 5 dB | 5 dB | 5 dB | 5 dB | 1 dB | 1 dB | 5 dB |
| | Max freq (kHz) and output (dB HL) | 16 (60 dB) | 16 (60 dB) | 12.5 (95 dB) | 16 (60 dB) | 12.5 (90 dB) | 14 (75 dB) | 16 (40 dB) |
| | Tinnitus Loudness (dB HL / SL /SPL) | 2 (dB SL and HL) | dB HL and dB SL | dB HL | dB HL | dB SPL | dB HL | dB HL |
| | Frequency steps | 50 Hz | 0.25–8 kHz: ½ octave; >8 kHz: ⅙ octave | 25 Hz | 1 Hz | 1 Hz | At PTA frequency | 10 Hz |
| | Loudness steps | 1 dB | 2 dB | 5 dB | 1 dB | 1 dB | 1dB | 5 dB |
| | Inserts | EARtone GOLD 3A (3M) | EARtone GOLD 3A (3M) | ER2 (Etymotic) | ER2 (Etymotic) | ER2 (Etymotic) | ER2 (Etymotic) | ER2 (Etymotic) |
| | Acoustic chamber | Yes | No | No | No | Yes | Yes | No |
| Electroencephalography | EEG system | EGI Philips (Magstim) | SynAmps 2 (Neuroscan) | ActiveTwo (BioSemi) | EEGoMylab (AntNeuro) | ActiChamp (Brainproducts) | ActiChamp (Brainproducts) | ActiChamp (Brainproducts) |
| | Electrodes | Passive | Passive | Active | Passive | Active | Active | Passive |
| | Electrode cap | Hydrocel GSN 64 (EGI) | Quik-Cap Neo Net 64 (Compumedics NeuroScan) | 64 (ECI) | Waveguard original cap 64ch 10/10 shildes tyco 68 (ANT-neuro) | actiCAP slim 64 (Brainproducts) | actiCAP slim 64 (Brainproducts) | Braincap TMS 64 (Brainproducts) |
| | Ref. Electrode | Cz | Between Cz and CPz | Cz | CPz | Fz | FCz | FCz |
| | Blink recording | Yes | Yes | No | No | Yes | No | No |
| | Line noise (Hz) | 60 | 60 | 50 | 50 | 50 | 50 | 50 |
| Presentation | OS | Win 10 | Win 10 | Win 10 | | Win 11 | Win 10 | Win 10 |
| | Psychopy version | 2021.2.3 | 2023.2.3 | 2021.2.3 | 2021.2.3 | 2022.2.5 | 2021.2.3 | 2021.2.3 |
| | Sound Calibration | CAL250 calibrator | B&K 4231 / CAL250 calibrator | GRAS 43AG Ear & Cheek Simulator | GRAS 43AG Ear & Cheek Simulator | B&K 4152 / sound level meter (NTi Audio XLR II) | B&K 4152 KEMAR | sound level meter (NTi Audio XLR) |
| | CPU | Intel Core i3-41060 3.60 GHz | Intel (R) Xeon (R) E-2146G 3.60 GHz | i5 3.0 GHz | Intel(R) Pentium(R) Gold G5400 3.70 GHz | i7 10th gen | AMD Ryzen 3 5300U 2.60 GHz | i7 2.9 GHz |
| | RAM | 4 GB | 32 GB | 16 GB | 4 GB | 16 GB | 8 GB | 16 GB |

**Table S2. Self-reported otological and medical history**

| Characteristic | Eligible pool (N = 562; 36 missing) | BBN Controls (n = 223; 34 missing) | BBN Cases (n = 232) |
|---|---|---|---|
| **Other diagnosed ear condition, n (%)[a]** | | | |
| Acoustic trauma | 18 (3.4) | 1 (0.5) | 15 (6.5) |
| Ear barotrauma | 3 (0.6) | 1 (0.5) | 2 (0.9) |
| Presbycusis | 18 (3.4) | 2 (1.1) | 15 (6.5) |
| Sudden hearing loss | 26 (4.9) | 2 (1.1) | 20 (8.6) |
| Other hearing loss | 23 (4.4) | 2 (1.1) | 13 (5.6) |
| Meniere's disease | 0 (0.0) | 0 (0.0) | 0 (0.0) |
| Acoustic neuroma | 0 (0.0) | 0 (0.0) | 0 (0.0) |
| Acute otitis | 25 (4.8) | 6 (3.2) | 13 (5.6) |
| Serous otitis / Eustachian tube dysfunction | 9 (1.7) | 2 (1.1) | 6 (2.6) |
| Chronic otitis | 6 (1.1) | 1 (0.5) | 4 (1.7) |
| Otosclerosis | 0 (0.0) | 0 (0.0) | 0 (0.0) |
| Other ear disorders | 12 (2.3) | 2 (1.1) | 9 (3.9) |
| No | 414 (78.7) | 174 (92.1) | 154 (66.4) |
| **Past procedures, n (%)[a]** | | | |
| Ear surgery | 18 (3.4) | 6 (3.2) | 10 (4.3) |
| Dental surgery | 220 (41.8) | 81 (42.9) | 99 (42.7) |
| Neurosurgery | 4 (0.8) | 4 (2.1) | 0 (0.0) |
| Lumbar puncture | 15 (2.9) | 5 (2.6) | 3 (1.3) |
| Chemotherapy | 8 (1.5) | 3 (1.6) | 4 (1.7) |
| Head and neck radiotherapy | 5 (1.0) | 1 (0.5) | 2 (0.9) |
| Electroconvulsive therapy | 7 (1.3) | 0 (0.0) | 6 (2.6) |
| Other procedure | 43 (8.2) | 14 (7.4) | 20 (8.6) |
| None of these | 266 (50.6) | 92 (48.7) | 114 (49.1) |
| **Pain syndromes, n (%)[a]** | | | |
| Headache | 73 (13.9) | 19 (10.1) | 44 (19.0) |
| Neck pain | 84 (16.0) | 19 (10.1) | 54 (23.3) |
| Ear pain | 9 (1.7) | 0 (0.0) | 8 (3.4) |
| Temporomandibular joint pain | 32 (6.1) | 8 (4.2) | 17 (7.3) |
| Pain in the face | 4 (0.8) | 0 (0.0) | 2 (0.9) |
| Other | 37 (7.0) | 6 (3.2) | 26 (11.2) |
| No | 371 (70.5) | 155 (82.0) | 133 (57.3) |
| **Clinician-diagnosed conditions, n (%)[a]** | | | |
| Temporomandibular joint disorder | 16 (3.0) | 2 (1.1) | 8 (3.4) |
| Dental problems | 30 (5.7) | 12 (6.3) | 11 (4.7) |
| Meningitis | 1 (0.2) | 1 (0.5) | 0 (0.0) |
| Multiple sclerosis | 0 (0.0) | 0 (0.0) | 0 (0.0) |
| Epilepsy | 1 (0.2) | 0 (0.0) | 1 (0.4) |
| Stroke | 1 (0.2) | 0 (0.0) | 1 (0.4) |
| Other cerebrovascular disease | 0 (0.0) | 0 (0.0) | 0 (0.0) |
| Dementia | 0 (0.0) | 0 (0.0) | 0 (0.0) |
| Other neurologic disease | 7 (1.3) | 2 (1.1) | 4 (1.7) |
| Anxiety | 38 (7.2) | 8 (4.2) | 20 (8.6) |
| Depression | 30 (5.7) | 11 (5.8) | 15 (6.5) |
| Emotional trauma | 14 (2.7) | 3 (1.6) | 8 (3.4) |
| Excessive stress | 19 (3.6) | 5 (2.6) | 11 (4.7) |
| Difficulty falling asleep | 53 (10.1) | 13 (6.9) | 31 (13.4) |
| Difficulty staying asleep | 44 (8.4) | 12 (6.3) | 26 (11.2) |
| Low blood pressure | 23 (4.4) | 8 (4.2) | 15 (6.5) |
| High blood pressure | 73 (13.9) | 27 (14.3) | 32 (13.8) |
| Myocardial infarction | 4 (0.8) | 1 (0.5) | 2 (0.9) |
| Thyroid disorder | 45 (8.6) | 19 (10.1) | 19 (8.2) |
| Diabetes | 14 (2.7) | 3 (1.6) | 7 (3.0) |
| Hyperinsulinemia | 2 (0.4) | 0 (0.0) | 2 (0.9) |
| Increased cholesterol | 63 (12.0) | 19 (10.1) | 32 (13.8) |
| Rheumatoid arthritis | 4 (0.8) | 2 (1.1) | 0 (0.0) |
| Systemic lupus erythematosus | 1 (0.2) | 0 (0.0) | 1 (0.4) |
| Chronic sinusitis | 6 (1.1) | 0 (0.0) | 5 (2.2) |
| Nasal septum deviation | 18 (3.4) | 5 (2.6) | 11 (4.7) |
| Syphilis | 1 (0.2) | 1 (0.5) | 0 (0.0) |
| HIV | 0 (0.0) | 0 (0.0) | 0 (0.0) |

| | | | |
|---|---|---|---|
| Lyme disease | 3 (0.6) | 0 (0.0) | 2 (0.9) |
| Anaemia | 8 (1.5) | 2 (1.1) | 3 (1.3) |
| Instability / balance disorders | 1 (0.2) | 1 (0.5) | 0 (0.0) |
| Acid/gastroesophageal reflux | 43 (8.2) | 17 (9.0) | 20 (8.6) |
| Globus hystericus | 2 (0.4) | 0 (0.0) | 2 (0.9) |
| Other | 26 (4.9) | 10 (5.3) | 11 (4.7) |
| None | 249 (47.3) | 106 (56.1) | 87 (37.5) |

a. Multiple-response (checkbox) questions, column percentages may sum to >100%. Percentages use item-specific non-missing denominators (n shown per item). Some participants did not complete every item (e.g., controls from Zurich did not answer these items). Eligible pool = all screened participants; BBN Controls/Cases, QC-passed non-tinnitus/tinnitus groups.

**Table S3. Recruitment, eligibility and data availability by site.**

| | UTA | UIUC | TCD | BCG | UZH | UKT | UR | All sites |
|---|---|---|---|---|---|---|---|---|
| Recruited via targeted mail | — | — | — | 226 | — | 225 | 186 | 637 |
| Recruited via flyers and adverts | 151 | 336 | 185 | — | 117 | — | 222 | 1,011 |
| Responded | 120 | 302 | 147 | 176 | — | 199 | 103 | 1,047 |
| Assessed for eligibility | 108 | 108 | 130 | 159 | 117 | 95 | 103 | 820 |
| Excluded at eligibility assessment | 25 | 25 | 43 | 76 | 30 | 14 | 23 | 236 |
| **Eligible participants** | **83** | **83** | **87** | **83** | **87** | **81** | **80** | **584** |
| Without a complete record in the final dataset | 1 | 0 | 4 | 6 | 9 | 2 | 0 | 22 |
| **Complete dataset available** | **82** | **83** | **83** | **77** | **78** | **79** | **80** | **562** |
| *of whom tinnitus cases* | 37 | 43 | 41 | 39 | 42 | 40 | 40 | 282 |
| *of whom controls* | 45 | 40 | 42 | 38 | 36 | 39 | 40 | 280 |

Site codes: UTA, UIUC, TCD, BCG, UZH, UKT, UR. Counts are individuals. Response counts were not recorded at UZH, so the total of 1,047 is the sum of the remaining six sites and the step from recruitment to eligibility assessment does not subtract exactly. Complete dataset available denotes a record present in the final merged dataset. It is a subset of the eligible sample and is not the denominator for any single-paradigm analysis, each of which draws on the eligible sample and applies its own quality control.

**Table S4. Reasons for exclusion at eligibility assessment, by site.**

| | UTA | UIUC | TCD | BCG | UZH | UKT | UR | All sites |
|---|---|---|---|---|---|---|---|---|
| Tinnitus not chronic | 4 | 4 | — | 4 | 10 | — | — | 22 |
| Age > 70 years | 6 | — | 2 | 5 | 3 | — | 3 | 19 |
| MoCA < 26 | 3 | 1 | — | — | — | 3 | — | 7 |
| HQ > 27[a] | 3 | 1 | 2 | 2 | — | 5 | 3 | 16 |
| Loudness discomfort level < 70 dB HL[b] | — | 2 | — | — | — | — | — | 2 |
| Objective tinnitus | — | — | — | — | 2 | — | — | 2 |
| Otosclerosis or acoustic neuroma | — | — | — | 3 | — | — | 4 | 7 |
| Ménière disease | — | — | 2 | 1 | — | — | 1 | 4 |
| Vestibular migraine | — | — | — | 2 | — | — | — | 2 |
| Serious internal, neurological or psychiatric condition | 1 | 1 | — | 6 | 2 | — | 1 | 11 |
| Epilepsy or other disorder of the central nervous system | — | — | 2 | 3 | 1 | — | — | 6 |
| Clinically relevant drug, medication or alcohol abuse | — | — | 1 | — | 1 | — | — | 2 |
| Severe hearing loss | — | 3 | 2 | 5 | 2 | — | 5 | 17 |
| Unilateral deafness | — | — | 1 | 2 | — | — | — | 3 |
| HADS > 14[c] | 3 | — | — | 1 | — | 3 | 1 | 8 |
| Not matchable to a counterpart | — | 5 | 24 | 18 | — | — | 1 | 48 |
| Lost interest, contact lost or illness | 5 | 8 | 7 | 24 | — | 1 | 4 | 49 |
| Reason not recorded | — | — | — | — | 9 | 2 | — | 11 |

a. HQ was administered at UZH using a 3-scale response in comparison to 4-scale in the other sites, therefore exclusion based on HQ could not be applied at UZH. b. Loudness discomfort Levels was only used at UIUC as part of clinical routine for exclusion purposes. c. HADS > 14 was initially used at several sites for exclusion and was modified during recruitment.

**Table S5. Audiometer and maximum output (dB HL) in the extended high-frequency range, by site.**

| Site | Audiometer | 8 kHz | 10 kHz | 11.2 kHz | 12.5 kHz | 14 kHz | 16 kHz |
|---|---|---|---|---|---|---|---|
| UTA | GSI Audiostar | 90 | 100 | n.c. | 90 | n.c. | 60 |
| UIUC | Equinox Interacoustics, HDA200 | — | 100 | 95 | 90 | 80 | 60 |
| TCD | Horentek Hekto audiometer, HDA 300 | 105 | 100 | 95 | 95 | n.c. | n.c. |
| BCG | Med Rx Studio | 105 | 100 | 90 | 70 | 70 | 60 |
| UZH | Multithreshold Matlab, RME UCX II, HDA200 | 90 | n.c. | n.c. | 90 | n.c. | n.c. |
| UKT | Auritec AT1000 | 100 | 95 | 95 | 85 | 75 | n.c. |
| UR | Astera 2 Otometrics, HDA 300 | 75 | 75 | 80 | 70 | 60 | 40 |
| **No-response entries recorded as maximum + 5 dB (ear measurements)** | | | | | | | |
| UTA | | 2 | 2 | — | 6 | — | 32 |
| UIUC | | — | 0 | 0 | 0 | 0 | 2 |
| TCD | | 0 | 0 | 0 | 0 | — | — |
| BCG | | 0 | 0 | 1 | 5 | 7 | 18 |
| UZH | | 0 | — | — | 28 | — | — |
| UKT | | 0 | 0 | 0 | 0 | 0 | — |
| UR | | 5 | 7 | 18 | 45 | 107 | 134 |

Values are the maximum output of the audiometer in use at that site, in dB HL. Frequencies marked not collected (n.c.) lie outside the range of that device and/or were not measured. Maximum output at a given frequency differs across sites by up to 30 dB at 8 kHz and 25 dB at 12.5 kHz. Where a participant did not respond at a site's maximum output, the threshold was recorded as that maximum plus 5 dB, the convention agreed across the consortium and standard in clinical screening. Those entries are no-response codes rather than measured thresholds, and are counted in the lower block. Extended high-frequency thresholds are therefore right-censored at a site-dependent level. No-response entries occur only at 8 kHz and above. None of the frequencies contributing to $PTA_4$ (0.5, 1, 2 and 4 kHz) is affected.

### Table S6. BBN inhibition ratio ROC AUCs

| | Apparent | | | LOSO-CV | | | Five-fold CV | | |
|---|---|---|---|---|---|---|---|---|---|
| **Model** | **nT** | **nC** | **AUC (95% CI)** | **nT** | **nC** | **AUC (95% CI)** | **nT** | **nC** | **AUC (95% CI)** |
| **— Full cohort —** | | | | | | | | | |
| Raw IR | 232 | 223 | 0.610 (0.559–0.660) | 227 | 220 | 0.592 (0.541–0.643) | 232 | 222 | 0.586 (0.534–0.637) |
| Raw IR + covariates | 232 | 223 | 0.672 (0.622–0.720) | 227 | 220 | 0.623 (0.573–0.671) | 232 | 222 | 0.636 (0.586–0.686) |
| Raw IR + covariates + pschy. | 221 | 220 | 0.700 (0.651–0.747) | 216 | 217 | 0.645 (0.594–0.695) | 221 | 219 | 0.658 (0.607–0.707) |
| Raw IR + covariates + 8 kHz | 227 | 223 | 0.699 (0.651–0.746) | 222 | 220 | 0.647 (0.597–0.695) | 227 | 222 | 0.662 (0.612–0.711) |
| Raw IR + covariates + 8 kHz + psych. | 221 | 220 | 0.729 (0.682–0.773) | 216 | 217 | 0.673 (0.626–0.720) | 221 | 219 | 0.687 (0.638–0.733) |
| **—Matched cohort —** | | | | | | | | | |
| Raw IR | 199 | 199 | 0.642 (0.587–0.696) | 195 | 197 | 0.615 (0.560–0.669) | 199 | 197 | 0.605 (0.549–0.661) |
| Raw IR + covariates | 199 | 199 | 0.663 (0.608–0.714) | 195 | 197 | 0.578 (0.530–0.626) | 199 | 197 | 0.614 (0.560–0.667) |
| Raw IR + covariates + pschy. | 192 | 197 | 0.701 (0.650–0.752) | 188 | 195 | 0.644 (0.590–0.697) | 192 | 195 | 0.652 (0.598–0.706) |
| Raw IR + covariates + 8 kHz | 198 | 199 | 0.701 (0.649–0.752) | 194 | 197 | 0.632 (0.579–0.684) | 198 | 197 | 0.664 (0.611–0.716) |
| Raw IR + covariates + 8 kHz + psych. | 192 | 197 | 0.741 (0.693–0.788) | 188 | 195 | 0.679 (0.624–0.732) | 192 | 195 | 0.695 (0.642–0.746) |

Covariates are Site, Age, Gender and $PTA_4$; pschy. are PSQ, HADS-A and HADS-D scores; 8 kHz, bilateral mean 8kHz hearing threshold. Confidence intervals are two-sided 95%.

### Table S7. Group difference in N100 inhibition in the broadband noise carrier — matched cohort.

| Site / fold | nT | nC | Δ mean, pp (95% CI) | p (unadjusted) | β, pp (95% CI) | p (adjusted) | FDR q | Adjusted d (95% CI) |
|---|---|---|---|---|---|---|---|---|
| **— Cluster-based group comparison —** | | | | | **Cluster-corrected permutation p = 0.0012** | | | |
| **UIUC** | 31 | 36 | −22.5 (−45.4 to 0.4) | 0.027 | −20.8 (−42.3 to 0.6) | 0.028 | 0.043 | −0.470 (−1.062 to 0.072) |
| **TCD** | 31 | 39 | −24.0 (−44.7 to −3.2) | 0.012 | −24.7 (−45.8 to −3.5) | 0.011 | 0.029 | −0.556 (−1.048 to −0.107) |
| **BCG** | 36 | 26 | −52.8 (−82.9 to −22.8) | <0.001 | −54.3 (−77.0 to −31.7) | <0.001 | <0.001 | −1.226 (−1.910 to −0.618) |
| **UZH** | 34 | 29 | −10.5 (−33.1 to 12.1) | 0.178 | −10.8 (−32.9 to 11.3) | 0.169 | 0.169 | −0.244 (−0.735 to 0.255) |
| **UKT** | 36 | 32 | −16.9 (−34.2 to 0.3) | 0.027 | −15.9 (−37.3 to 5.4) | 0.072 | 0.086 | −0.359 (−0.773 to 0.014) |
| **UR** | 31 | 37 | −25.3 (−42.3 to −8.3) | 0.002 | −23.7 (−45.1 to −2.4) | 0.015 | 0.029 | −0.536 (−0.955 to −0.158) |
| **All sites** | 199 | 199 | −25.4 (−34.3 to −16.6) | <0.001 | −24.6 (−33.5 to −15.7) | <0.001 | — | −0.552 (−0.749 to −0.367) |
| **— Site influence: site omission sensitivity —** | | | | | | | | |
| **omit UIUC** | 168 | 163 | −26.0 (−35.6 to −16.3) | <0.001 | −25.3 (−35.0 to −15.7) | <0.001 | <0.001 | −0.575 (−0.790 to −0.378) |
| **omit TCD** | 168 | 160 | −26.2 (−36.0 to −16.3) | <0.001 | −24.4 (−34.2 to −14.6) | <0.001 | <0.001 | −0.544 (−0.768 to −0.340) |
| **omit BCG** | 163 | 173 | −19.9 (−28.8 to −11.0) | <0.001 | −19.2 (−28.1 to −10.3) | <0.001 | <0.001 | −0.468 (−0.689 to −0.260) |
| **omit UZH** | 165 | 170 | −28.1 (−37.7 to −18.4) | <0.001 | −27.3 (−37.1 to −17.6) | <0.001 | <0.001 | −0.615 (−0.833 to −0.417) |
| **omit UKT** | 163 | 167 | −27.4 (−37.4 to −17.3) | <0.001 | −26.6 (−36.7 to −16.5) | <0.001 | <0.001 | −0.575 (−0.794 to −0.380) |
| **omit UR** | 168 | 162 | −25.0 (−35.1 to −14.9) | <0.001 | −24.7 (−34.9 to −14.4) | <0.001 | <0.001 | −0.531 (−0.748 to −0.327) |
| **— LOSO cross-validation: held-out site —** | | | | | | | | |
| **UIUC** | 31 | 36 | −23.2 (−46.2 to −0.3) | 0.024 | −21.9 (−45.6 to 1.7) | 0.034 | 0.052 | −0.453 (−0.946 to 0.050) |
| **TCD** | 31 | 39 | −21.2 (−43.9 to 1.5) | 0.033 | −21.6 (−44.5 to 1.2) | 0.032 | 0.052 | −0.454 (−0.927 to 0.008) |
| **BCG** | 33 | 24 | −33.4 (−66.4 to −0.4) | 0.024 | −33.9 (−66.9 to −0.8) | 0.022 | 0.052 | −0.551 (−1.047 to −0.068) |
| **UZH** | 34 | 29 | −16.3 (−41.4 to 8.8) | 0.100 | −16.9 (−42.1 to 8.4) | 0.093 | 0.093 | −0.338 (−0.824 to 0.135) |
| **UKT** | 36 | 32 | −20.0 (−42.3 to 2.2) | 0.038 | −20.6 (−43.0 to 1.7) | 0.035 | 0.052 | −0.448 (−0.812 to −0.023) |
| **UR** | 30 | 37 | −17.8 (−41.7 to 6.2) | 0.072 | −16.6 (−40.6 to 7.4) | 0.086 | 0.093 | −0.340 (−1.030 to 0.134) |
| **— Five-fold cross-validation: held-out fold —** | | | | | | | | |
| **Fold 1** | 40 | 40 | −20.3 (−42.5 to 1.8) | 0.036 | −17.9 (−40.1 to 4.4) | 0.057 | 0.057 | −0.357 (−0.753 to 0.040) |
| **Fold 2** | 40 | 40 | −27.6 (−53.5 to −1.7) | 0.019 | −26.6 (−52.2 to −0.9) | 0.021 | 0.034 | −0.462 (−0.879 to −0.035) |
| **Fold 3** | 40 | 39 | −24.5 (−49.2 to 0.1) | 0.026 | −24.7 (−49.1 to −0.2) | 0.024 | 0.034 | −0.453 (−0.898 to −0.010) |
| **Fold 4** | 40 | 39 | −20.9 (−43.1 to 1.4) | 0.033 | −21.9 (−44.1 to 0.4) | 0.027 | 0.034 | −0.441 (−0.876 to −0.006) |
| **Fold 5** | 39 | 39 | −31.5 (−59.9 to −3.1) | 0.015 | −31.7 (−60.4 to −3.1) | 0.015 | 0.034 | −0.499 (−0.953 to −0.052) |

Δ mean is the unadjusted difference in inhibition ratio between tinnitus and control participants, in percentage points; negative values indicate reduced inhibition in tinnitus. β is the covariate-adjusted group coefficient from ordinary least squares (×100, percentage points), adjusted for Age, Gender and $PTA_4$ within each site and additionally for Site in the pooled rows. Adjusted d is β divided by the model residual root-mean-square error; it is not a pooled-standard-deviation Cohen's d. All p values are one-sided in the pre-specified direction (tinnitus < control); confidence intervals are two-sided 95%, so an interval may include zero where $p < 0.05$. FDR q values are Benjamini–Hochberg adjusted p values computed separately within each block of this table; the pooled row is not a member of the per-site family and carries no q. The cluster-corrected permutation p for the pooled cluster-based contrast is given in the block header. Cross-validation rows re-derive cluster selection and the within-group outlier fence from training data alone, so a held-out participant contributes no score either because no electrode in the training-selected cluster meets the amplitude criterion or because their inhibition ratio falls outside the training-derived fence. These removed 0 and 6 participants respectively under leave-one-site-out (392 analysed) and 1 and 1 under five-fold cross-validation (396 analysed), from 398 entering each block.

**Table S8. Desikan–Killiany (DK) parcel composition of the two full-cohort TFCE-significant source clusters containing more than 100 vertices.**

| Rank | DK parcel | Hemisphere | Vertices | % Distribution |
|---|---|---|---|---|
| ***Cluster 1:*** *1,616 vertices; minimum TFCE-corrected vertex p = 0.0022* | | | | |
| 1 | precuneus | RH | 196 | 11.0 |
| 2 | superiortemporal | RH | 196 | 11.0 |
| 3 | insula | RH | 171 | 9.6 |
| 4 | middletemporal | RH | 169 | 9.5 |
| 5 | fusiform | RH | 143 | 8.0 |
| 6 | inferiortemporal | RH | 135 | 7.6 |
| 7 | bankssts | RH | 92 | 5.2 |
| 8 | parahippocampal | RH | 80 | 4.5 |
| 9 | isthmuscingulate | RH | 74 | 4.2 |
| 10 | lingual | RH | 29 | 1.6 |
| 11 | entorhinal | RH | 23 | 1.3 |
| 12 | supramarginal | RH | 21 | 1.2 |
| 13 | transversetemporal | RH | 17 | 1.0 |
| 14 | temporalpole | RH | 14 | 0.8 |
| 15 | inferiorparietal | RH | 4 | 0.2 |
| 16 | lateralorbitofrontal | RH | 3 | 0.2 |
| 17 | parsopercularis | RH | 2 | 0.1 |
| 18 | parstriangularis | RH | 2 | 0.1 |
| 19 | postcentral | RH | 2 | 0.1 |
| 20 | precentral | RH | 1 | 0.1 |
| 21 | superiorparietal | RH | 1 | 0.1 |
| | unassigned/non-DK | RH | 241 | 13.5 |
| ***Cluster 2:*** *165 vertices; minimum TFCE-corrected vertex p = 0.0262* | | | | |
| 1 | superiorparietal | RH | 155 | 8.7 |
| 2 | precuneus | RH | 9 | 0.5 |
| 3 | cuneus | RH | 1 | 0.1 |

Each vertex was assigned to a DK cortical parcel. The table reports the rank, DK parcel, hemisphere, and number of significant vertices within each parcel. Parcels are ordered by decreasing vertex count and LH and RH indicate the left and right hemispheres, respectively. "unassigned/non-DK" vertices do not correspond to a named DK parcel. The minimum TFCE p-value represents the smallest corrected vertex-wise p-value within each cluster.

### Supplemental Figures

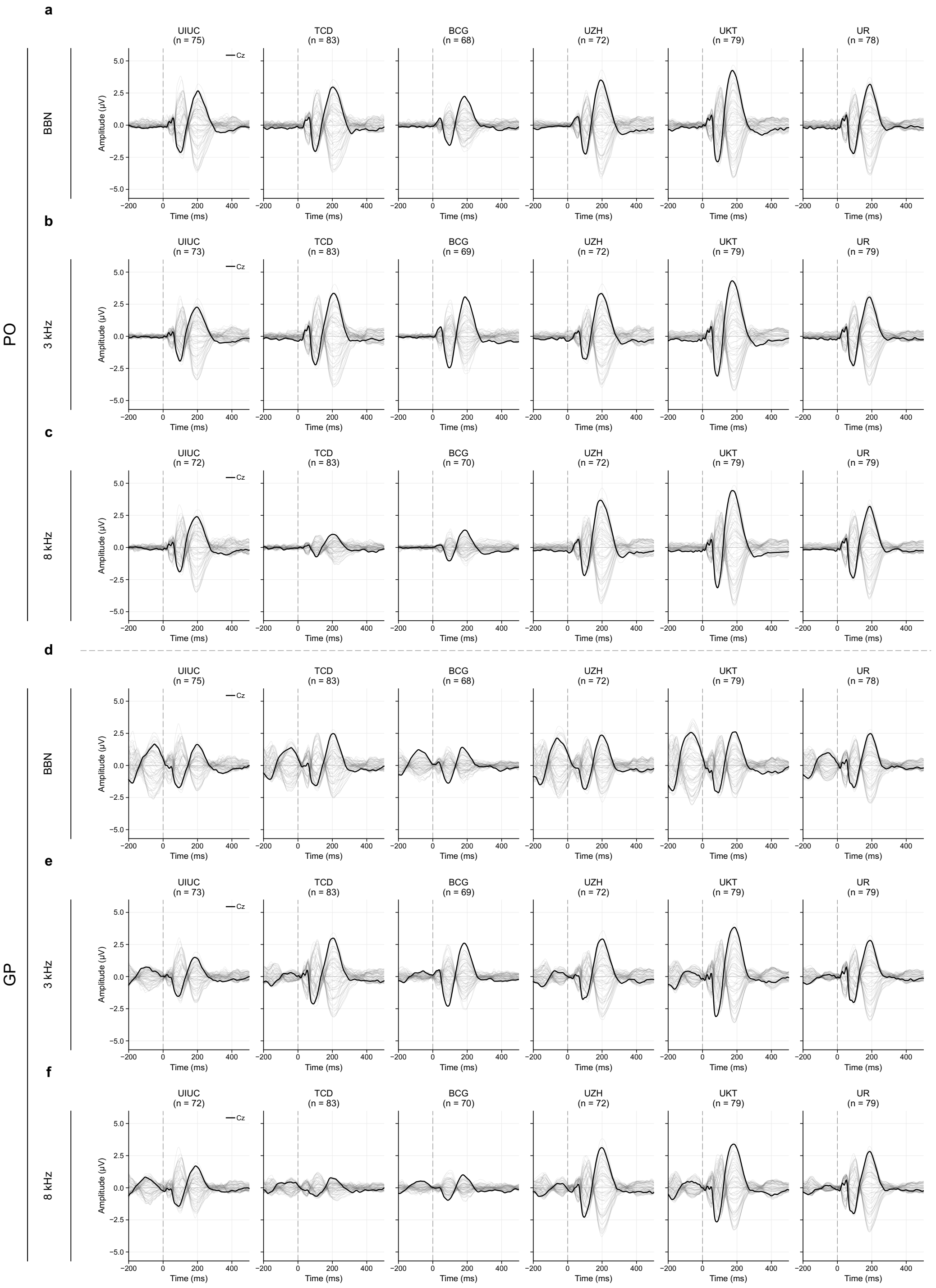

**Figure S1. Grand-average auditory evoked responses across recording sites and carrier conditions.** Butterfly plots show grand-average event-related potentials for pulse-only (PO) responses to (a) BBN, (b) 3 kHz, and (c) 8 kHz carriers, and corresponding gap–pulse (GP) responses (d–f). Columns represent the six recording sites. Traces show all 64 EEG channels, with Cz highlighted in black and the remaining channels in gray. Time is expressed relative to pulse onset, indicated by the vertical dashed line. All panels share the same amplitude scale. Tinnitus participants and controls were pooled within each site. Each carrier-specific cohort included 455 participants: 232 tinnitus participants and 223 controls for BBN and 8 kHz, and 229 tinnitus participants and 226 controls for 3 kHz. Site-specific sample sizes are indicated above each plot. Responses were averaged after preprocessing and session-level quality control.

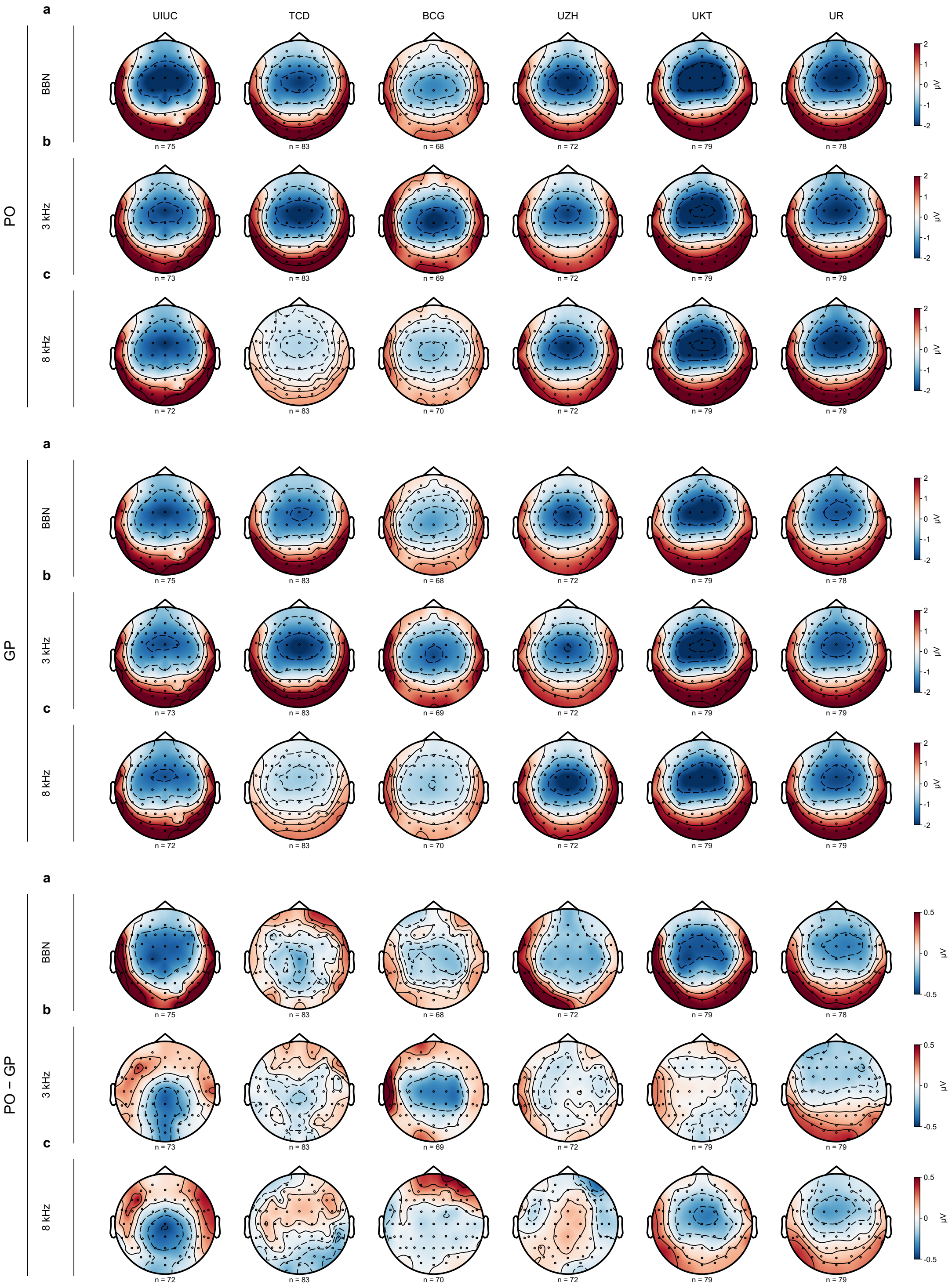

**Figure S2. N100 scalp topographies across recording sites and carrier conditions.** Scalp maps show grand-average signed EEG amplitudes over 80–120 ms after pulse onset. Within each block, (a) BBN, (b) 3 kHz, and (c) 8 kHz responses are shown across the six recording sites. The upper, middle, and lower blocks display PO responses, GP responses, and the signed PO minus GP amplitude difference, respectively. Black dots indicate electrode positions. Red and blue represent positive and negative amplitudes, respectively. Color scales range from −2 to +2 µV for PO and GP responses and from −0.5

to +0.5 μV for difference maps. Tinnitus participants and controls were pooled within each site. Each carrier included 455 participants, with site-specific sample sizes shown below the maps. These descriptive amplitude maps were generated after preprocessing and session-level quality control.

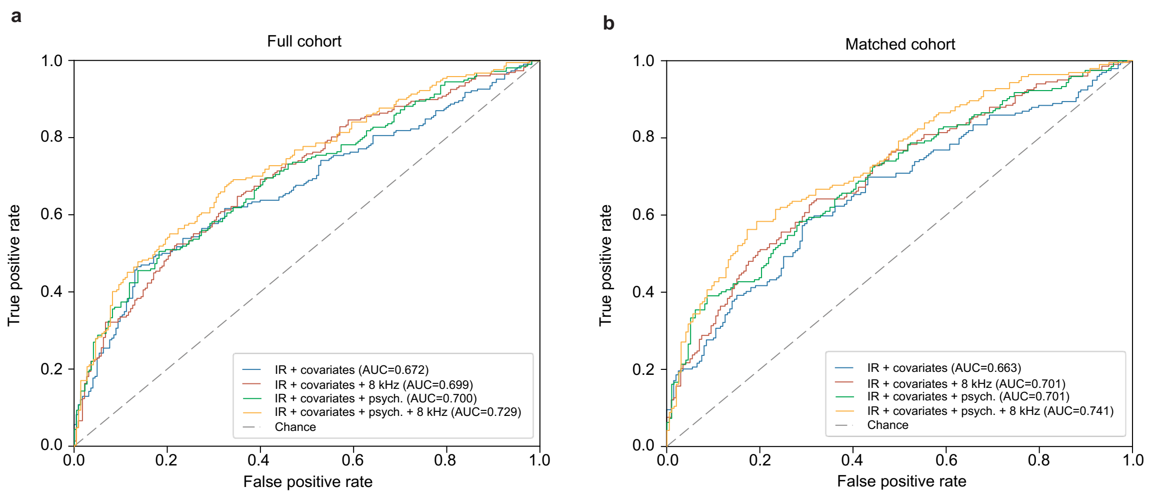

**Figure S3. ROC analysis of tinnitus classification using IR and additional clinical variables.** ROC curves comparing models combining IR with covariates (age, gender, $PTA_4$, and site), 8 kHz hearing threshold, and psychological measures (PSQ, HADS-A, and HADS-D). (a) Full cohort and (b) matched cohort. The area under the curve (AUC) is reported for each model.

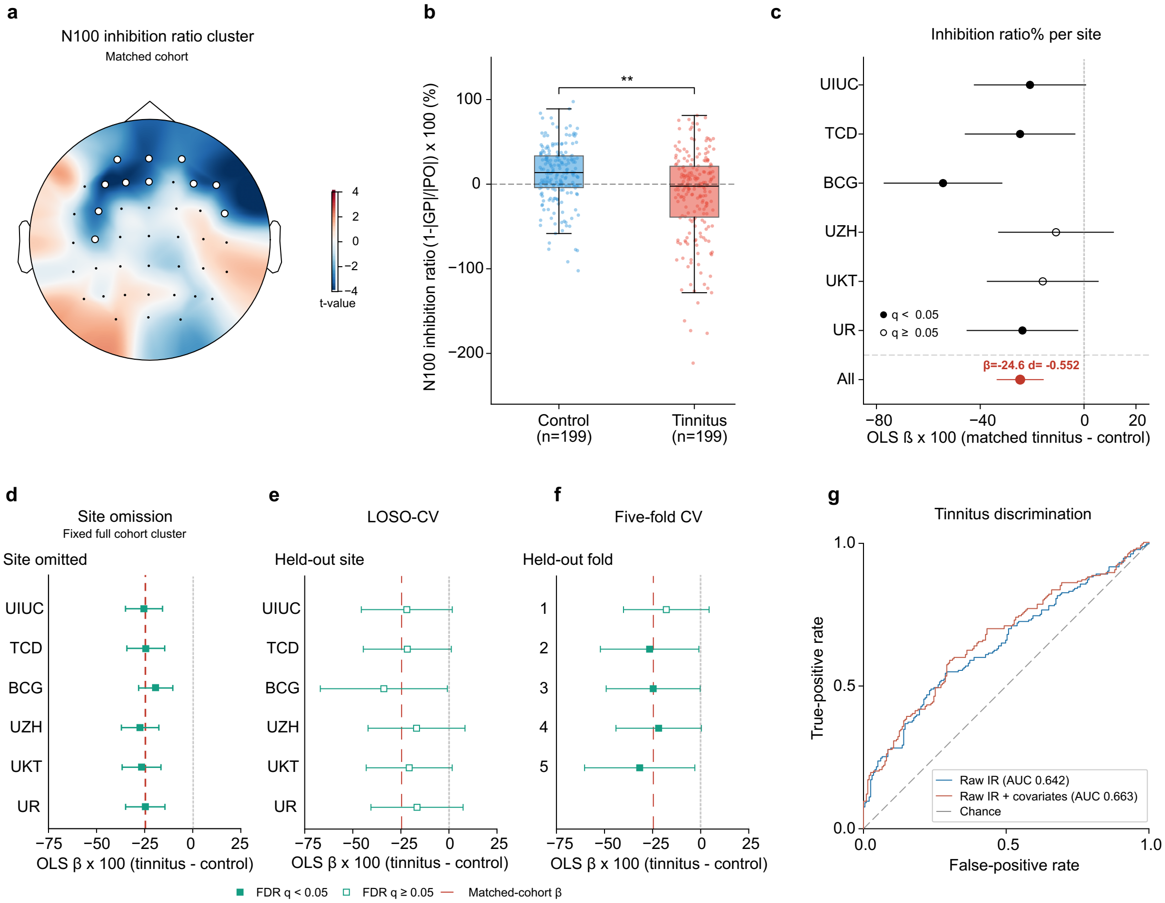

**Figure S4. BBN N100 inhibition and validation analyses in the matched cohort.** (a) Scalp map of t statistics for the tinnitus and control difference in BBN N100 inhibition ratio. White circles mark the 11-electrode cluster identified by covariate-adjusted (age, gender, $PTA_4$, and site) cluster-permutation testing in the matched cohort (AF3, AF4, AFz, C3, F1, F3, F4, F6, FC3, FC6, and Fz). The color scale represents the t values. (b) Covariate-adjusted participant inhibition ratios averaged across eligible cluster electrodes. Blue and red indicate controls and tinnitus participants, respectively. Points represent individual observations and boxplots summarize the group distributions. The bracket and asterisks indicate spatially corrected cluster-permutation significance (** $p < 0.01$). (c) Site-specific and pooled (red) adjusted OLS estimates of the tinnitus minus control difference in BBN N100 inhibition ratio. Horizontal lines represent 95% confidence intervals. Filled markers indicate BH-FDR $q < 0.05$ across the six site comparisons; open markers indicate $q \geq 0.05$. (d) Site omission analysis, excluding each site in turn while retaining the matched-cohort cluster. (e,f) Held-out estimates from LOSO-CV and five-fold CV, respectively, using training-derived N100 windows, cluster selection, and nuisance adjustment. Forest plots show β in percentage points and two-sided 95% OLS confidence intervals. Filled markers indicate BH-FDR $q < 0.05$ within each panel's comparison family; open markers indicate $q \geq 0.05$. Red dashed lines in panels d–f indicate the matched-cohort coefficient. (g) ROC curves for raw IR and IR combined with covariates (age, gender, $PTA_4$, and site).

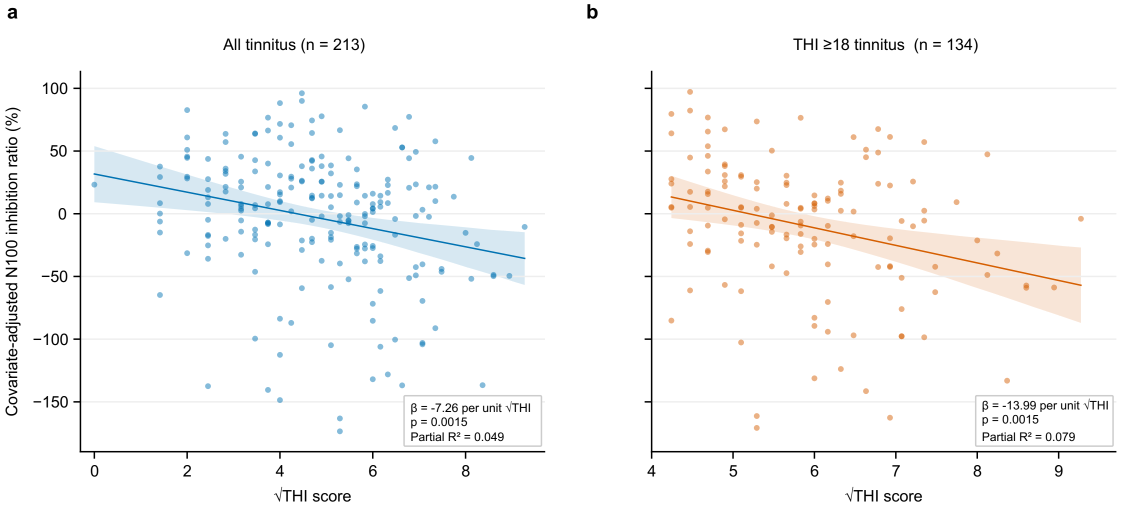

**Figure S5. Tinnitus severity correlates with the degree of impaired inhibition.** Covariate-adjusted associations between square-root of Tinnitus Handicap Inventory scores (√THI) and N100 inhibition ratio in (a) all eligible tinnitus participants (n = 213, blue) and (b) participants with THI ≥18 (n = 134; orange). Inhibition ratios were calculated from the fixed 11-electrode full cohort cluster. Points represent individual participants adjusted for age, gender, $PTA_4$, and site. Solid lines indicate OLS regression fits and shaded bands show 95% confidence intervals for the adjusted mean. Regression coefficients (β) express the change in inhibition ratio, in percentage points, per unit increase in √THI. Partial $R^2$ indicates the contribution of √THI after covariate adjustment. P-values are two-sided OLS values.

**STROBE statement — checklist for case–control studies**

Line numbers refer to the continuous line numbering of the submitted, line-numbered PDFs. References to the main manuscript are given as line numbers of that file (lines 1–1003). Where an item is addressed across a whole subsection, the line range of that subsection is given.

| Section / topic | Item | Recommendation (abbreviated) | Reported on line(s) |
|---|---|---|---|
| **Title and abstract** | **1a** | Study design indicated in the title or the abstract. | Title, lines 1–2; Abstract, line 49 ("a prospective, multicentre case–control study at seven sites in the United States and Europe"). |
| | **1b** | Informative and balanced summary of what was done and found. | Abstract, lines 47–64. |
| **Introduction** | **2** | Scientific background and rationale. | Introduction, lines 67–108. |
| | **3** | Specific objectives, including any prespecified hypotheses. | Introduction, lines 67–108; Results, lines 168–199 (prespecified narrowband hypothesis and its reversal). Methods, lines 440–444 — the protocol designated no primary or secondary outcome measures and no statistical analysis plan; the group contrast in each carrier and the diagnostic accuracy of the broadband contrast are designated here as the primary analyses, and all other analyses as exploratory and hypothesis-generating. |
| **Methods** | **4** | Key elements of study design, presented early. | Abstract, line 49; Methods, "Study Planning and Conduct", lines 423–432 (line 424, "TIDE is a prospective, multicentre case–control study"). |
| | **5** | Setting, locations and relevant dates. | Methods, lines 423–432 (seven clinical sites, September 2024 – October 2025). |
| | **6a** | Eligibility criteria; sources and methods of case ascertainment and control selection; rationale for the choice of cases and controls. | Methods, "Participants", lines 452–492; "Reference standard", lines 524–540. |
| | **6b** | For matched studies, matching criteria and the number of controls per case. | Methods, lines 665–666 (199 tinnitus–control pairs matched by gender, within five years of age and 5 dB PTA4; one control per case); Results, lines 210–211. |

| | | | |
|---|---|---|---|
| | **7** | Clearly defined outcomes, exposures, predictors, potential confounders and effect modifiers. | Methods, “Primary Outcome”, lines 541–543; “Reference standard”, lines 524–540; “Statistical Analysis”, lines 616–628. |
| | **8** | For each variable, sources of data and details of methods of assessment; comparability of assessment methods across groups. | Methods, lines 493–523 (questionnaires and audiometry) and 543–584 (EEG design, paradigm and preprocessing); Supplementary Materials, Tables S1 and S5. |
| | **9** | Efforts to address potential sources of bias. | Methods, lines 602–603 (post-hoc SNR threshold, stated as not pre-specified); lines 641–644 (circularity of cluster selection, with the cross-validation analyses given as the less circular estimate); lines 621–622 (outlier fence derived from training data only and applied unchanged to held-out participants); line 585 (analyst blinded to group); lines 598–600 (channel-agnostic SNR measure chosen to avoid circularity); Discussion, lines 379–396. Methods, lines 440–444 — primary versus exploratory designation, as at item 3. |
| | **10** | How the study size was arrived at. | Methods, “Ethics, registration and study size”, lines 433–451 (no a priori power calculation, line 444; target of 40 cases and 40 controls per site, line 447; the erroneous power statement in the published protocol disclosed and a correction requested, line 449; detectable effect size in the analysed sample, d = 0.233 at 80% power, lines 450–451). |
| | **11** | Explain how quantitative variables were handled; which groupings were chosen and why. | Methods, lines 617–620 (inhibition ratio, PO amplitude criterion > 0.1 μV, 3×IQR Tukey fence); lines 626–628 (AIC selected the linear form for both, so the reported model is linear in age and PTA4) (functional form of age and PTA4 compared as linear and natural cubic spline, chosen by AIC, which selected the linear form for both); |

| | | | |
|---|---|---|---|
| | | | lines 678–686 (rationale for the square-root THI transformation, with skewness, Shapiro–Wilk and leverage values before and after, and the disclosure that it was not pre-specified). |
| | **12a** | All statistical methods, including those used to control for confounding. | Methods, lines 616–628 (covariate-adjusted OLS with the fitted equation at line 625, adjusted for Age, Gender, PTA4 and Site); lines 629–644 (Freedman–Lane cluster permutation); lines 645–649 (cross-validation and Benjamini–Hochberg FDR within each family). Methods, lines 440–444 — primary versus exploratory designation, as at item 3. |
| | **12b** | Methods used to examine subgroups and interactions. | Results, lines 209–267 (matched cohort, severity regression, source reconstruction); Methods, lines 665–707. |
| | **12c** | How missing data were addressed. | Methods, line 504 (missing PSQ entries; ESIT-SQ not answered by controls at one site); lines 603–605 (recordings failing quality control treated as indeterminate, neither reclassified nor imputed); lines 606–615 (per-carrier counts after quality control, the carrier overlap, and the removal by listwise deletion of two participants without a derivable PTA4, 457 → 455); lines 622–623 (“All models were fitted on complete cases and no values were imputed”). |
| | **12d** | How matching of cases and controls was addressed. | Methods, lines 665–673 (cluster selection repeated within the matched cohort; matched pairs kept within the same fold; percentile-bootstrap resampling of matched pairs). |
| | **12e** | Sensitivity analyses. | Results, lines 168–230 (site omission, leave-one-site-out and five-fold cross-validation, matched cohort); Methods, lines 645–673; the SNR ≥ 1.5 sensitivity analysis is at lines 605–606. |

| | | | |
|---|---|---|---|
| **Results** | **13a** | Numbers of individuals at each stage of the study. | Results, lines 121–128; Methods, lines 606–615; Figure 1c, lines 940–955; Supplementary Materials, Table S3. |
| | **13b** | Reasons for non-participation at each stage. | Results, lines 123–125; Supplementary Materials, Table S4; Figure 1c, lines 940–955. |
| | **13c** | Consider use of a flow diagram. | Figure 1c, lines 940–955. The diagram carries recruitment, eligibility, site-level exclusion, per-carrier quality control and the 457 → 455 listwise-deletion step. The legend states that quality control was applied per carrier, so the three analysed samples differ in composition — 449 participants passed all three carriers and 461 passed at least one (lines 951–953) — and gives the cross-validation denominators, 447 for leave-one-site-out and 454 for five-fold (lines 953–955). The same denominators appear in the footnotes to Table 2 (lines 933–935) and in the footnote to Table S7. |
| | **14a** | Characteristics of study participants and information on exposures and potential confounders. | Table 1, lines 919–925. |
| | **14b** | Number of participants with missing data for each variable of interest. | Table 1 footnotes, lines 922–925; Supplementary Materials, Table S2 column headers; Methods, line 504. |
| | **15** | Report numbers in each exposure category, or summary measures of exposure. | Table 1, lines 919–925; Table 2, lines 928–935; Supplementary Materials, Table S7. |
| | **16a** | Unadjusted and confounder-adjusted estimates and their precision; which confounders were adjusted for and why. | Table 2, lines 928–935, and Supplementary Materials, Table S7 — each row carries the unadjusted Δ mean with 95% CI and p, and the adjusted β with 95% CI, p and FDR-adjusted q; the adjustment set is given at line 625 and in the Table 2 footnote, lines 930–932. |
| | **16b** | Category boundaries when continuous variables were categorized. | Methods, lines 501–504 (THI severity bands); Results, lines 236–239; the THI ≥ 18 subgroup is defined at line 678. |

| | | | |
|---|---|---|---|
| | **16c** | Translation of relative risk into absolute risk for a meaningful time period. | Not applicable — the outcome is a continuous inhibition ratio, not a risk. |
| | **17** | Other analyses done — subgroups, interactions and sensitivity analyses. | Results, lines 209–267 (matched cohort, severity regression, vertex-wise source reconstruction); Methods, lines 665–707; Figure 4, lines 993–1004; Supplementary Materials, Figures S3–S5 and Tables S6–S8. |
| **Discussion** | **18** | Summarise key results with reference to study objectives. | Discussion, lines 268–284. |
| | **19** | Limitations, sources of potential bias or imprecision, with direction and magnitude discussed. | Discussion, lines 379–406 (multi-site heterogeneity; circularity of cluster selection; under-representation of severe tinnitus and its link to the hyperacusis exclusion, with the direction of the resulting bias stated, line 395). |
| | **20** | Cautious overall interpretation considering objectives, limitations, multiplicity, and other evidence. | Discussion, lines 268–284 and 378–407; multiplicity is stated at line 388 ("Analyses across carriers, electrode clusters, sites, folds and source vertices were not corrected as a single family …") and the exploratory results are named there as hypothesis-generating; a further statement is in the Table 1 footnote, line 925. Methods, lines 440–444 — primary versus exploratory designation, as at item 3. |
| | **21** | Generalisability (external validity). | Discussion, lines 379–406; comparison with the companion resting-state and oddball paradigms, lines 397–406. |
| **Other information** | **22** | Funding sources and the role of the funders. | Acknowledgements, lines 729–736 (funding sources); Role of the Funder/Sponsor, lines 748–753; competing interests, lines 737–747; data availability, lines 709–713; code availability, lines 714–716. |

**STARD 2015 — checklist for studies of diagnostic accuracy**

The index test is reported as a continuous, covariate-adjusted score. No positivity threshold was defined and no participant was classified, so the items that depend on a threshold (12a in part, and 23) are addressed by that explicit statement rather than by a 2×2 table. This is stated in the Methods at lines 647–650.

| Section / topic | Item | Recommendation (abbreviated) | Reported on line(s) |
|---|---|---|---|
| **Title or abstract** | **1** | Identification as a study of diagnostic accuracy using at least one measure of accuracy. | Title, lines 1–2; Abstract, lines 58–59 (unadjusted AUC 0.610, 95% CI 0.559–0.660). |
| **Abstract** | **2** | Structured summary of design, methods, results and conclusions. | Abstract, lines 47–64. |
| **Introduction** | **3** | Scientific and clinical background, including the intended use and clinical role of the index test. | Introduction, lines 67–108; Methods, lines 423–424; Discussion, lines 397–406. |
| | **4** | Study objectives and hypotheses. | Introduction, lines 67–108; Results, lines 168–208. Methods, lines 440–444 — the protocol designated no primary or secondary outcome measures and no statistical analysis plan; the group contrast in each carrier and the diagnostic accuracy of the broadband contrast are designated here as the primary analyses, and all other analyses as exploratory and hypothesis-generating. |
| **Methods** | **5** | Whether data collection was planned before the index test and reference standard were performed. | Methods, line 424 (“TIDE is a prospective, multicentre case–control study”); registration before enrolment, lines 439–441. Methods, lines 440–444 — primary versus exploratory designation, as at item 4. |
| **Participants** | **6** | Eligibility criteria. | Methods, lines 452–492; Supplementary Materials, Table S4. |
| | **7** | On what basis potentially eligible participants were identified. | Methods, lines 453–455; Supplementary Materials, Table S3; Figure 1c, lines 940–955. |
| | **8** | Where and when potentially eligible participants were identified. | Methods, lines 423–432 (seven sites, September 2024 – October 2025); Results, lines 121–123. |
| | **9** | Whether participants formed a consecutive, random or convenience series. | Methods, line 455 (“Participants therefore formed a convenience series rather than a consecutive or random sample of the clinic populations”). |
| **Test methods** | **10a** | The index test, in sufficient detail to allow replication. | Methods, lines 544–573 (EEG design, paradigm, stimuli and recording) and |

| | | | |
|---|---|---|---|
| | | | 573–584 (preprocessing); Figures 1a–b, lines 940–949; Figure 2, lines 957–975; code repository, lines 714–716. |
| | **10b** | The reference standard, in sufficient detail to allow replication. | Methods, lines 524–540. |
| | **11** | Rationale for choosing the reference standard. | Methods, lines 536–540 (no objective confirmatory test exists; the consequences of reference-standard misclassification for attainable accuracy are stated). |
| | **12a** | Definition of and rationale for index-test positivity cut-offs, and whether prespecified or exploratory. | Methods, line 655 — “No positivity threshold was defined: the index test is reported as a continuous covariate-adjusted score, and no classification of individual participants was performed.” |
| | **12b** | Definition of and rationale for reference-standard positivity cut-offs. | Methods, lines 524–536 (case and control definitions); lines 501–504 (THI severity bands). |
| | **13a** | Whether clinical information and reference-standard results were available to the index-test performers or readers. | Methods, line 585 (“The analyst was blinded to group”). |
| | **13b** | Whether clinical information and index-test results were available to the assessors of the reference standard. | Methods, lines 524–535 — case and control status was established at the eligibility interview, before any EEG was recorded, so index-test results were not available to the assessors. |
| **Analysis** | **14** | Methods for estimating or comparing measures of diagnostic accuracy. | Methods, lines 650–664 (ROC construction on covariate-adjusted scores, leave-one-site-out and five-fold cross-validation, 20,000-resample conditional bootstrap confidence intervals preserving site and group counts); matched cohort, lines 665–673. Methods, lines 440–444 — primary versus exploratory designation, as at item 4. |
| | **15** | How indeterminate index-test or reference-standard results were handled. | Methods, lines 602–605 — recordings failing the SNR criterion were treated as indeterminate index-test results, were neither reclassified nor imputed, and contributed to no analysis. |
| | **16** | How missing data on the index test and reference standard were handled. | Methods, lines 606–615 and lines 622–623 (complete-case analysis, no imputation); line 504 (questionnaire missingness). |

| | | | |
|---|---|---|---|
| | **17** | Any analyses of variability in diagnostic accuracy, and how they were handled. | Results, lines 168–208 (site-level, site-omission and cross-validated estimates) and 208–229 (matched cohort); Table 2, lines 928–935; Supplementary Materials, Tables S6 and S7; Figure 3d–g, lines 976–992; Figures S3 and S4. |
| | **18** | Intended sample size and how it was determined. | Methods, lines 433–451. |
| **Results** | **19** | Flow of participants, using a diagram. | Figure 1c, lines 940–955; Supplementary Materials, Table S3; Methods, lines 606–615. The legend gives the carrier overlap (449 passed all three carriers, 461 at least one; lines 951–953) and the cross-validation denominators (447 leave-one-site-out, 454 five-fold; lines 953–955), which also appear in the footnotes to Table 2 (lines 933–935) and to Supplementary Table S7. |
| | **20** | Baseline demographic and clinical characteristics of participants. | Table 1, lines 919–925; Supplementary Materials, Table S2. |
| | **21a** | Distribution of severity of disease in those with the target condition. | Results, lines 236–239 — of the 232 tinnitus participants analysed for the broadband carrier, THI fell in the negligible (0–16) band for 84 (36.2%), mild (18–36) for 92 (39.7%), moderate (38–56) for 45 (19.4%), severe (58–76) for 9 (3.9%) and catastrophic (78–100) for 2 (0.9%); Table 1, lines 919–925. |
| | **21b** | Distribution of alternative diagnoses in those without the target condition. | Methods, lines 469–478; Supplementary Materials, Table S2 (self-reported otological and medical history, reported separately for controls and cases). |
| | **22** | Time interval and any clinical interventions between index test and reference standard. | Methods, lines 524–535 — both within the same study visit, the reference standard first, with no intervening intervention. |
| **Test results** | **23** | Cross-tabulation of the index-test results by the results of the reference standard. | Not applicable — no positivity threshold was defined and no participant was classified (Methods, line 655). Discrimination is reported as AUC with confidence intervals instead (item 24). |

| | | | |
|---|---|---|---|
| | **24** | Estimates of diagnostic accuracy and their precision. | Results, lines 201–208 — full cohort apparent AUC 0.610 (95% CI 0.559–0.660) for the raw inhibition ratio, 0.672 (0.622–0.720) with Site, Age, Gender and PTA4, and 0.729 (0.682–0.773) with the 8 kHz threshold and psychological measures added; matched cohort, lines 225–230 (0.642, 95% CI 0.587–0.696 for the raw ratio). Held-out estimates for every model are in Supplementary Table S6 (full cohort raw ratio: leave-one-site-out 0.592, 95% CI 0.541–0.643; five-fold 0.586, 0.534–0.637). Figure 3g, lines 976–992; Figures S3 and S4g. |
| | **25** | Any adverse events from performing the index test or the reference standard. | Results, lines 127–128 — no adverse events related to the paradigm were reported; participant comfort and sleepiness were checked via intercom after each block. Methods, line 572. |
| **Discussion** | **26** | Study limitations, including sources of potential bias, statistical uncertainty and generalisability. | Discussion, lines 379–406; line 388 (multiplicity); Methods, lines 536–540 (reference-standard misclassification as a bound on attainable accuracy). |
| | **27** | Implications for practice, including the intended use and clinical role of the index test. | Discussion, lines 268–284 and 396–407. |
| **Other information** | **28** | Registration number and name of the registry. | Methods, lines 439–440 — ClinicalTrials.gov NCT06520865; Abstract, line 61. |
| | **29** | Where the full study protocol can be accessed. | Methods, line 432 (the protocol is published and cited); the correction requested to its power statement is disclosed at line 449. |
| | **30** | Sources of funding and other support; role of the funders. | Acknowledgements, lines 729–736; Role of the Funder/Sponsor, lines 748–753. |

**Note on two items reported with deliberate limits**

Item STARD 12a / 23 — no positivity threshold. The inhibition ratio is reported as a continuous covariate-adjusted score. Defining a cut-off on these data would be exploratory and would be derived from the same sample used to estimate accuracy, so no threshold was set, no participant was classified, and no 2×2 table, sensitivity or specificity is reported. The limited individual discrimination this reflects is the manuscript's central negative finding.

Item STROBE 12a — functional form of the covariates. Age and PTA4 were each compared as a linear term and as a natural cubic spline, with the form chosen by AIC, which selected the linear form for both (lines 618–622). The ΔAIC values themselves are not tabulated.
